# Impact of the COVID-19 pandemic on low back pain management in commercially insured and Medicare Advantage cohorts. A retrospective cohort study

**DOI:** 10.1101/2023.05.15.23289993

**Authors:** David Elton, Meng Zhang

## Abstract

**Background:** The COVID-19 (COVID) pandemic has been associated with care seeking and delivery system changes. Before COVID the management of low back pain (LBP) was variable and a common source of low-value care. The purpose of this retrospective cohort study was to examine how the management of LBP changed during the COVID pandemic in commercially insured (CI) and Medicare Advantage (MA) cohorts.

**Methods:** A US national sample of LBP episodes with a duration of less than 91 days experienced during 2019-2021 was analyzed. Independent variables included whether an individual had CI or MA coverage, and the timing of LBP onset. Secondary independent variables included individual’s home address State. Dependent measures included the percent of individuals initially contacting eighteen types of health care provider (HCP) and receiving twenty-two types of health care services, and total episode cost. Early and late COVID measures were compared with a pre-COVID baseline to examine COVID related change. The impact of the stringency of State level COVID public policy response was evaluated.

**Results:** The study included 222,043 CI and 466,125 MA complete episodes of LBP. During the pre-COVID period the MA cohort was older (MA median 72 vs. CI 45), with higher percent female (61% vs. 52%), and from zip codes with a higher Area Deprivation Index (median 52 vs. CI 38). MA and CI cohort attributes remained nearly identical in the early and late COVID periods.

Initial contact with licensed acupuncturists (LAc risk ratio (RR) 0.66) and physical therapists (PT RR 0.82) in the CI cohort, and with PTs (RR 0.78), urgent care (RR 0.86), and emergency medicine (RR 0.87) in the MA cohort experienced the largest decreases during the early COVID period. The largest increase in the CI cohort was to PCPs (RR 1.08), and in the MA cohort to PCPs (RR 1.11) and nurse practitioners (RR 1.09). During the late COVID period the largest decreases in the CI cohort were to neurologists (RR 0.84), PTs (RR 0.86), and physical medicine and rehabilitation physicians (RR PMR 0.87) and in the MA cohort to rheumatologists (RR 0.81), PMR (RR 0.89) and pain management physicians (RR 0.89). The largest increases during the late COVID period in the CI cohort were to radiologists (RR 1.22), hospitals (RR 1.07) and orthopedic surgeons (OS RR 1.04) and in the MA cohort to LAcs (RR 1.32), radiologists (RR 1.11) and hospitals (RR 1.10).

Compared to the pre-COVID period during the early COVID period the percent of episodes including most health care services was unchanged or reduced. In the late COVID period in both the CI and MA cohorts the percent of episodes with imaging studies increased, MRI (RR CI 1.15, MA 1.21), CT (RR 1.16, 1.16), and plain film radiology (RR 1.06, 1.06).

The stringency of State COVID public policy responses was not associated with significant variability in either the type of HCP initially contacted, or services received for LBP.

**Conclusions:** In both CI and MA LBP cohorts COVID was associated with changes in the types of HCP initially contacted and subsequent services provided. Guideline concordant first-line service use declined during COVID, and the rate of diagnostic imaging was higher in the late COVID period than the pre-COVID period.

## Background

Low back pain (LBP) prevalence, disability, and costs are well understood.^1–4^ Less well-understood is how the management of LBP was impacted by health care delivery system disruption caused by and variable public policy responses to ^5–7^ COVID-19 (COVID), declared a global pandemic on March 11, 2020 by the World Health Organization (WHO).^8–11^

High quality LBP clinical practice guidelines (CPG) emphasize natural history, self-care, and non-pharmaceutical services as first-line approaches.^12–14^ While LBP CPGs have been widely available for many years, LBP has been estimated to be a source of almost half of low-value spending in the US with low-value defined as costs incurred with little to no associated benefit.^15, 16^

Variation in LBP utilization and cost outcomes have been analyzed using the type of health care provider (HCP) initially contacted and subsequent care pathways.^17–19^ Primary care providers (PCP) and chiropractors (DC) are the most common types of HCP initially contacted by an individual with LBP.^19^ LBP is the second most common reason for visiting a PCP.^20^ Early access to DCs, physical therapists (PT), or licensed acupuncturists (LAc) is associated with lower rates of low-value services like advanced spinal imaging studies and lower rates of opioid prescriptions.^19, 21–24^

There is a growing body of research exploring how the COVID pandemic impacted the management of LBP. A study of fifteen low-value and sixteen high-value services found that during the early COVID surge from April to June of 2020 the rate of use of all services decreased, except for opioid use for back and neck pain, and headache.^25^ Similarly, compared to a 2019 pre-COVID baseline, in the first four months of the COVID pandemic non-pharmaceutical service use decreased and prescription opioid use increased for back, neck or extremity pain.^26^ In Canada, individuals with pain reported increasing pharmaceutical intake to compensate for loss of availability of non-pharmaceutical options.^27^ Reductions in PCP visits, emergency department visits, interventional procedures, and neurosurgical consultations have all been observed for LBP during the COVID pandemic.^28–33^

Non-pharmaceutical services like osteopathic manipulative treatment, active care, and manual therapy have been reported to have declined, particularly early in the COVID pandemic.^34–36^ Telehealth alternatives to in-office non-pharmaceutical services were made available and an increase in the use was observed early in the pandemic.^37, 38^ Telehealth alternatives have been found to be generally effective, however there are signs of declining use and a return to in-office delivery of non-pharmaceutical services.^39–44^

The aim of this study was to explore how the COVID pandemic impacted care-seeking for, and the management of, LBP in commercially insured (CI) and Medicare Advantage (MA) cohorts. The hypothesis was during the COVID pandemic a lower percentage of individuals with LBP sought initial treatment from HCPs like DCs, PTs and LAcs, and that this was associated with an increase in the rate of prescription pharmaceutical use.

## Methods

### Study design, population, setting and data sources

This is a retrospective cohort study of individuals seen by one or more HCPs for a complete episode of LBP. Individuals were from a single national insurer administering CI and MA benefit plans. An enrollee database included de-identified enrollment records, and administrative claims data for all inpatient and outpatient services, and pharmacy prescriptions. De-identified in and out-of-network HCP demographic information and professional licensure status were available in an HCP database. ZIP code level 2020 population race and ethnicity data was obtained from the US Census Bureau,^45^ 2019 adjusted gross income (AGI) data from the Internal Revenue Service,^46^ and 2020 socioeconomic Area Deprivation Index (ADI) data from the University of Wisconsin Neighborhood Atlas^®^ database.^47, 48^

With data being de-identified or a Limited Data Set in compliance with the Health Insurance Portability and Accountability Act and customer requirements, the UnitedHealth Group Office of Human Research Affairs Institutional Review Board determined that this study was exempt from ongoing Institutional Review Boards oversight. The study was conducted and reported based on the Strengthening the Reporting of Observational Studies in Epidemiology (STROBE) guidelines [Supplement – STROBE Checklist].^49^

The impact of numerous unmeasurable and unknown confounders, and confounders of measurable hypothesized confounders were likely amplified by the impact of the COVID pandemic and variable public policy responses.^6, 7^ [Supplement – Public Policy] As an alternative to adopting the standard practice of using potentially inadequate approaches such as propensity score matching^50^ to control for available yet incomplete potential confounders derived from administrative claims data to attempt to generate causal insights^51, 52^, the study was designed to address a literature gap of presenting actual, unadjusted associations between individual demographic attributes, HCP selection, and episodic characteristics for the management of LBP during the COVID pandemic. These confounders, significant cohort differences, insurance benefit design differences, and a different distribution of episodes among States resulted in an inability to directly compare CI and MA measures, and no such comparison should be attempted with study data.

### Unit of analysis and cohort selection

Episode of care has been shown to be a valid way to organize administrative claims data to summarize care pathways and analyze the rate and timing of use of services provided for a condition.^19, 53^ The Symmetry^®^ Episode Treatment Groups^®^ (ETG^®^) and Episode Risk Groups^®^ (ERG^®^) version 9.5 methodologies and definitions were used to translate administrative claims data into discrete episodes of care.^54^ A complete episode was defined as having at least 91-day pre- and 61-day post-episode clean periods, during which no services were provided by any HCP for any LBP diagnosis. The episode duration was the number of days between the first and last date of service for an episode.

The cohort consisted of individuals aged 18 years and older with a complete episode of LBP commencing and ending during the calendar years 2019-2021. To align with the timing of the WHO pandemic declaration^8–11^, the pre-COVID period was defined as episodes starting between 3/1/2019 and 2/29/2020, the early COVID period was 3/1/2020 to 2/28/2021, and the late COVID period was 3/1/2021 to 1/28/2022.

Episodes starting in the pre-COVID period had up to 35 months of post-onset duration potential, episodes starting in the early COVID period had up to 23 months and episodes starting in the late COVID period had up to 11 months. The resulting different episode volumes and duration distributions in the pre, early and late COVID periods and within and between the CI and MA cohorts was a potential confounder of a variety of study variables. As one example, MA episode durations were approximately double CI durations indicating a greater prevalence of chronic LBP in the MA cohort. To address this potential study limitation, a sensitivity analysis was conducted to evaluate limiting the cohort to episodes with a duration of less than 61 or less than 91 days. Based on the sensitivity analysis, the study was limited to episodes with a duration of less than 91 days. This approach balanced episode volume across pre, early and late COVID periods, preserved at least a 14-day median episode duration, and reduced or eliminated the original significant difference in episode duration between pre, early and late COVID periods and between the CI and MI cohorts. This approach may have partially addressed differing LBP clinical complexity in CI and MA cohorts and reduced but did not eliminate episodes crossing the pre, early and post COVID measurement periods. [Supplement – Episode Duration] Results for the full cohort are available as supplemental tables. [Supplement – Table 1 – Full Cohort] [Supplement – Table 2 – Full Cohort] [Supplement – Table 3 – Full Cohort]

**Table 1.** Cohort, population and episode attributes for complete low back pain episodes <91 days duration

|  |  |  |  |
| --- | --- | --- | --- |
|  | Pre-COVID | Early COVID | Late COVID |
| Commercial Insurance |  |  |  |
| Episodes | 67158 | 78050 | 76835 |
| Individuals | 65019 | 75987 | 76341 |
| # Unique HCPs | 45360 | 52494 | 52138 |
| Total Cost | \$60,367,712 | \$67,940,430 | \$65,098,792 |
| Individual Attributes - % or Median (Q1, Q3) |  |  |  |
| % Female | 51.7% | 51.1% | 51.6% |
| Age | 45 (32, 55) | 45 (33, 55) | 46 (33, 56) |
| ERG® Risk Score | 1.1 (0.5, 2.3) | 1.1 (0.5, 2.5) | 1.3 (0.6, 2.7) |
| Individual Home Address 5-digit Zip Code Attributes - Median (Q1, Q3) |  |  |  |
| % Non-Hispanic White (NHW) | 66.1% (47.0%, 79.2%) | 66.4% (47.3%, 79.6%) | 66.2% (46.5%, 79.4%) |
| Area Deprivation Index (ADI) | 38 (19, 58) | 40 (21, 60) | 39 (20, 59) |
| Household Adjusted Gross Income (AGI) - 1000s | 76 (56, 113) | 74 (55, 109) | 75 (55, 111) |
| Total Episode Attributes - Median (Q1, Q3) |  |  |  |
| Cost | \$245 (92, 698) | \$220 (70, 640) | \$250 (81, 704) |
| # Healthcare Providers Seen | 1 (1, 2) | 1 (1, 2) | 1 (1, 2) |
| Duration (days) | 12 (1, 43) | 11 (1, 43) | 13 (1, 44) |
| Medicare Advantage |  |  |  |
| Episodes | 141258 | 152406 | 172461 |
| Individuals | 136941 | 148540 | 171799 |
| # Unique HCPs | 75594 | 82075 | 90360 |
| Total Cost | \$63,369,127 | \$65,486,484 | \$82,097,246 |
| Individual Attributes - % or Median (Q1, Q3) |  |  |  |
| % Female | 60.7% | 60.1% | 60.4% |
| Age | 72 (67, 78) | 72 (67, 78) | 73 (68, 78) |
| ERG® Risk Score | 0.9 (0.4, 1.7) | 1.0 (0.5, 1.8) | 1.1 (0.5, 1.9) |
| Individual Home Address 5-digit Zip Code Attributes - Median (Q1, Q3) |  |  |  |
| % Non-Hispanic White (NHW) | 68.0% (45.9%, 81.7%) | 68.6% (46.8%, 81.9%) | 69.0% (47.8%, 82.1%) |
| Area Deprivation Index (ADI) | 52 (33, 70) | 53 (34, 71) | 53 (34, 70) |
| Household Adjusted Gross Income (AGI) - 1000s | 62 (49, 84) | 61 (49, 82) | 62 (50, 83) |
| Total Episode Attributes - Median (Q1, Q3) |  |  |  |
| Cost | \$131 (35, 398) | \$114 (30, 360) | \$147 (36, 445) |
| # Healthcare Providers Seen | 1 (1, 2) | 1 (1, 2) | 1 (1, 2) |
| Duration (days) | 15 (1, 49) | 13 (1, 49) | 16 (1, 50) |

Within CI and MA, compared to Pre-COVID baseline, cells in red are not significantly different - Mann-Whitney U test (p=0.05)
Comparing MA to CI, all MA measures were significantly different from CI measures except # of Healthcare Providers Seen - Mann-Whitney U test (p=0.05)

**Table 2.** Type of healthcare provider initially contacted for low back pain - duration <91 days

|  |  | % of episodes including service |  |  | RR (95% CI) Compared to Pre-COVID |  |
| --- | --- | --- | --- | --- | --- | --- |
|  |  | Pre-COVID | Early COVID | Late COVID | Early COVID | Late COVID |
| Commercial Insurance (CI) |  |  |  |  |  |  |
| Episodes |  | 67158 | 78050 | 76835 |  |  |
| Non-Rx | Doctor of Chiropractic (DC) | 26.6% | 26.8% | 25.4% | 1.01 (0.99, 1.02) | 0.95 (0.94, 0.97) |
|  | Physical Therapist (PT) | 3.8% | 3.1% | 3.3% | 0.82 (0.77, 0.86) | 0.86 (0.82, 0.91) |
|  | Licensed Acupuncturist (LAC) | 0.2% | 0.2% | 0.2% | 0.66 (0.52, 0.83) | 0.84 (0.68, 1.05) |
| Primary Care | Primary Care Physician (PCP) | 20.4% | 22.0% | 19.7% | 1.08 (1.06, 1.10) | 0.97 (0.95, 0.99) |
|  | Nurse Practitioner (Nurse) | 5.6% | 5.8% | 5.7% | 1.04 (1.00, 1.08) | 1.02 (0.98, 1.07) |
|  | Physician Assistant (PA) | 5.1% | 4.7% | 4.8% | 0.93 (0.89, 0.98) | 0.95 (0.91, 1.00) |
|  | Doctor of Osteopathy (DO) | 0.2% | 0.2% | 0.2% | 1.04 (0.83, 1.30) | 0.94 (0.75, 1.18) |
| Physician Specialist | Orthopedic Surgeon (OS) | 8.5% | 8.5% | 8.9% | 1.00 (0.96, 1.03) | 1.04 (1.00, 1.07) |
|  | Physical Medicine & Rehabilitation (PMR) | 3.2% | 2.8% | 2.8% | 0.87 (0.82, 0.92) | 0.87 (0.82, 0.93) |
|  | Pain Management (PM) | 1.7% | 1.8% | 1.7% | 1.01 (0.94, 1.09) | 0.97 (0.90, 1.05) |
|  | Neurologist (Neuro) | 0.5% | 0.5% | 0.4% | 0.99 (0.86, 1.15) | 0.84 (0.72, 0.98) |
|  | Rheumatologist (Rheum) | 0.5% | 0.5% | 0.5% | 1.05 (0.90, 1.22) | 1.05 (0.90, 1.22) |
|  | Neurosurgeon (NS) | 0.3% | 0.3% | 0.4% | 1.00 (0.84, 1.20) | 1.07 (0.90, 1.28) |
|  | Other Physician (Oth) | 1.4% | 1.8% | 1.6% | 1.28 (1.18, 1.39) | 1.14 (1.05, 1.24) |
| Emergency Medicine/<br>Urgent Care | Hospital (Hosp) | 10.2% | 9.5% | 11.0% | 0.93 (0.90, 0.96) | 1.07 (1.04, 1.11) |
|  | Radiologist (Rad) | 8.2% | 8.2% | 10.0% | 1.01 (0.97, 1.04) | 1.22 (1.18, 1.26) |
|  | Emergency Medicine (EM) | 2.6% | 2.5% | 2.6% | 0.97 (0.91, 1.03) | 1.02 (0.96, 1.09) |
|  | Urgent Care (UC) | 0.9% | 0.8% | 0.8% | 0.89 (0.80, 0.99) | 0.90 (0.81, 1.01) |
| Medicare Advantage (MA) |  |  |  |  |  |  |
| Episodes |  | 141258 | 152406 | 172461 |  |  |
| Non-Rx | Doctor of Chiropractic (DC) | 9.6% | 8.6% | 9.1% | 0.90 (0.88, 0.92) | 0.95 (0.93, 0.97) |
|  | Physical Therapist (PT) | 3.5% | 2.7% | 3.3% | 0.78 (0.75, 0.81) | 0.94 (0.90, 0.98) |
|  | Licensed Acupuncturist (LAC) | 0.1% | 0.1% | 0.1% | 0.81 (0.62, 1.06) | 1.32 (1.05, 1.66) |
| Primary Care | Primary Care Physician (PCP) | 31.5% | 35.0% | 29.5% | 1.11 (1.10, 1.12) | 0.94 (0.93, 0.95) |
|  | Nurse Practitioner (Nurse) | 6.1% | 6.7% | 6.2% | 1.09 (1.06, 1.12) | 1.01 (0.98, 1.04) |
|  | Physician Assistant (PA) | 4.5% | 4.4% | 4.7% | 0.99 (0.96, 1.03) | 1.04 (1.01, 1.08) |
|  | Doctor of Osteopathy (DO) | 0.1% | 0.1% | 0.1% | 0.86 (0.70, 1.05) | 1.01 (0.83, 1.22) |
| Physician Specialist | Orthopedic Surgeon (OS) | 6.5% | 6.4% | 6.8% | 0.99 (0.96, 1.02) | 1.05 (1.02, 1.08) |
|  | Physical Medicine & Rehabilitation (PMR) | 2.3% | 2.0% | 2.1% | 0.87 (0.82, 0.91) | 0.89 (0.85, 0.94) |
|  | Pain Management (PM) | 2.9% | 2.7% | 2.6% | 0.91 (0.88, 0.95) | 0.89 (0.85, 0.92) |
|  | Neurologist (Neuro) | 0.5% | 0.6% | 0.5% | 1.10 (1.00, 1.21) | 0.94 (0.85, 1.03) |
|  | Rheumatologist (Rheum) | 0.8% | 0.7% | 0.6% | 0.93 (0.86, 1.01) | 0.81 (0.75, 0.88) |
|  | Neurosurgeon (NS) | 0.3% | 0.3% | 0.3% | 0.91 (0.79, 1.04) | 0.88 (0.77, 1.01) |
|  | Other Physician (Oth) | 1.8% | 2.3% | 1.9% | 1.23 (1.17, 1.30) | 1.06 (1.01, 1.12) |
| Emergency Medicine/<br>Urgent Care | Hospital (Hosp) | 15.4% | 14.3% | 17.0% | 0.92 (0.91, 0.94) | 1.10 (1.08, 1.12) |
|  | Radiologist (Rad) | 11.6% | 11.2% | 12.9% | 0.96 (0.94, 0.98) | 1.11 (1.09, 1.13) |
|  | Emergency Medicine (EM) | 1.9% | 1.7% | 1.9% | 0.87 (0.82, 0.91) | 0.96 (0.91, 1.01) |
|  | Urgent Care (UC) | 0.5% | 0.4% | 0.4% | 0.86 (0.77, 0.96) | 0.91 (0.82, 1.01) |

Pre=Pre-COVID period, Early=first 12 months post-COVID, Late=13-24 months post-COVID
RR=Risk Ratio, CI=Confidence Interval
For risk ratio, cells in red indicate the confidence interval includes 1 and likelihood of HCP being initially contacted is not different than the pre-COVID reference

**Table 3.**
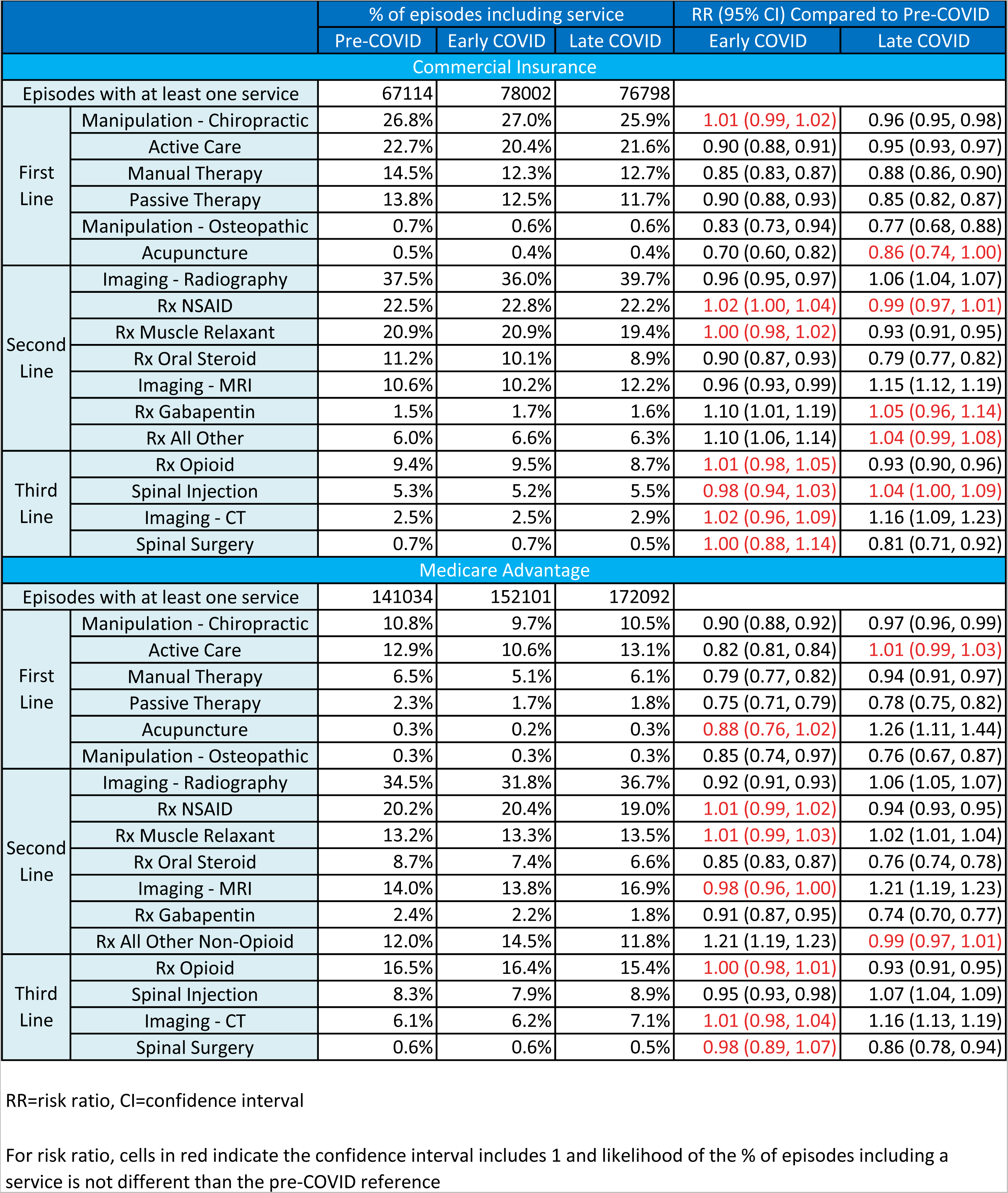
Type of healthcare services provided for low back pain with duration <91 days

Individuals with diagnoses of malignant and non-malignant neoplasms, fractures and other spinal trauma, infection, congenital deformities and scoliosis, autoimmune disorders, osteoporosis, and advanced arthritis were excluded from the analysis. This approach was used to address potential confounders associated the COVID pandemic impacting care seeking behaviors of individuals with a more complex LBP condition differently from individuals with less complex LBP.

### Variables

Data preprocessing, table generation, and initial analyses were performed in Python (Python Language Reference, Version 3.7.5., n.d.). A goodness-of-fit analysis was performed using D’Agostino’s K-squared test. Non-normally distributed data were reported using the median, interquartile range (IQR), quartile 1 (Q1), and quartile 3 (Q3). Where utilized, p-values do not control for the false discovery rate. Due to the tendency for odds ratios to exaggerate risk in situations where an outcome is relatively common, and as a measure more widely understood in associational analyses, risk ratios (RR) and associated 95% confidence intervals were reported.^55^

The primary independent variables were the individual’s timing of LBP onset and type of insurance coverage. The primary dependent variables were the percent of individuals with LBP initially contacting one of eighteen types of HCP, and the percent of episodes including twenty-two types of health care services. The main secondary dependent variable was the total cost of care for all reimbursed services provided by any HCP during an episode. Total episode cost included costs associated with all services provided for LBP during an episode, including those not specifically identified in the service categories used in the analyses. Costs for services for which an insurance claim was not submitted, and indirect costs associated with missed days at work or reduced productivity, were not available. Additional secondary dependent variables included episode duration and the number of different HCPs seen during an episode. Due to numerous confounders no attempt was made to calculate the prevalence of LBP during the pre, early and late COVID time periods.

To explore the impact of the level of stringency of State public policy responses on dependent variables Oxford Stringency Scores on 4/1/2020 and 4/1/2021 were used to rank order States from highest to lowest stringency, with a score of 1 indicating the highest stringency level.^7^ 4/1/2020 was selected as this was the first day of a month with complete rankings immediately following the March 11, 2020 global pandemic declaration. 4/1/2021 was selected to examine stringency in the late COVID period. States were categorized as being in the Top 25 (T25) most stringent States on both dates or the Lowest 25 (B25) on both dates. States that changed from T25 to B25 or B25 to T25 were categorized as Mixed. [Supplement – Public Policy]

## Results

The study included 688,168 complete episodes of LBP, with 222,043 CI and 466,125 MA episodes. Depending on the pre/early/late COVID measurement period, the CI cohort was 51-52% female with a median age of 45-46, and the MA cohort was 60-61% female with a median age of 72-73. While the scaling for the ERG® Risk Score, a measure of illness burden, was different for CI and MA cohorts, within each cohort the median ERG® Risk Score was constant across pre/early/late COVID periods. 5-digit zip code population attributes were also different for CI and MA cohorts and within a cohort were constant across pre/early/late COVID periods for each cohort. For CI median ADI was 38-40, AGI was $74k-$76k, and the % NHW was 66%. For MA median ADI was 52-53, AGI was $61k-$62k and % NHW was 68%-69%. Compared to the pre-COVID baseline, total episode cost decreased in the early COVID period, and increased in the late COVID period. Episode duration median and IQR was similar across time periods in both the CI and MA cohorts. Episodes were from all 50 States; however, this was not a US representative sample and the distribution of episodes among States was different in CI and MA cohorts. [Supplement – States] [Table 1]

During the pre-COVID period the CI cohort with LBP initially contacted DCs (26.6% of episodes) and PCPs (20.4%) most commonly. Hospitals (10.2%), orthopedic surgeons (OS 8.5%), radiologists (8.2%), nurse practitioners (5.6%), and physician assistants (PA 5.1%) were the only other HCP types seen initially for greater than 5% of episodes. [Figure 1] In the MA cohort during the pre-COVID period, PCPs (31.5%), hospitals (15.4%), and radiologists (11.6%) were most common. DCs (9.6), OS (6.5%) and nurses (6.1%) were the only other HCP types seen initially by greater than 5% of episodes in the MA cohort. [Figure 2] [Table 2]

**Figure 1.**
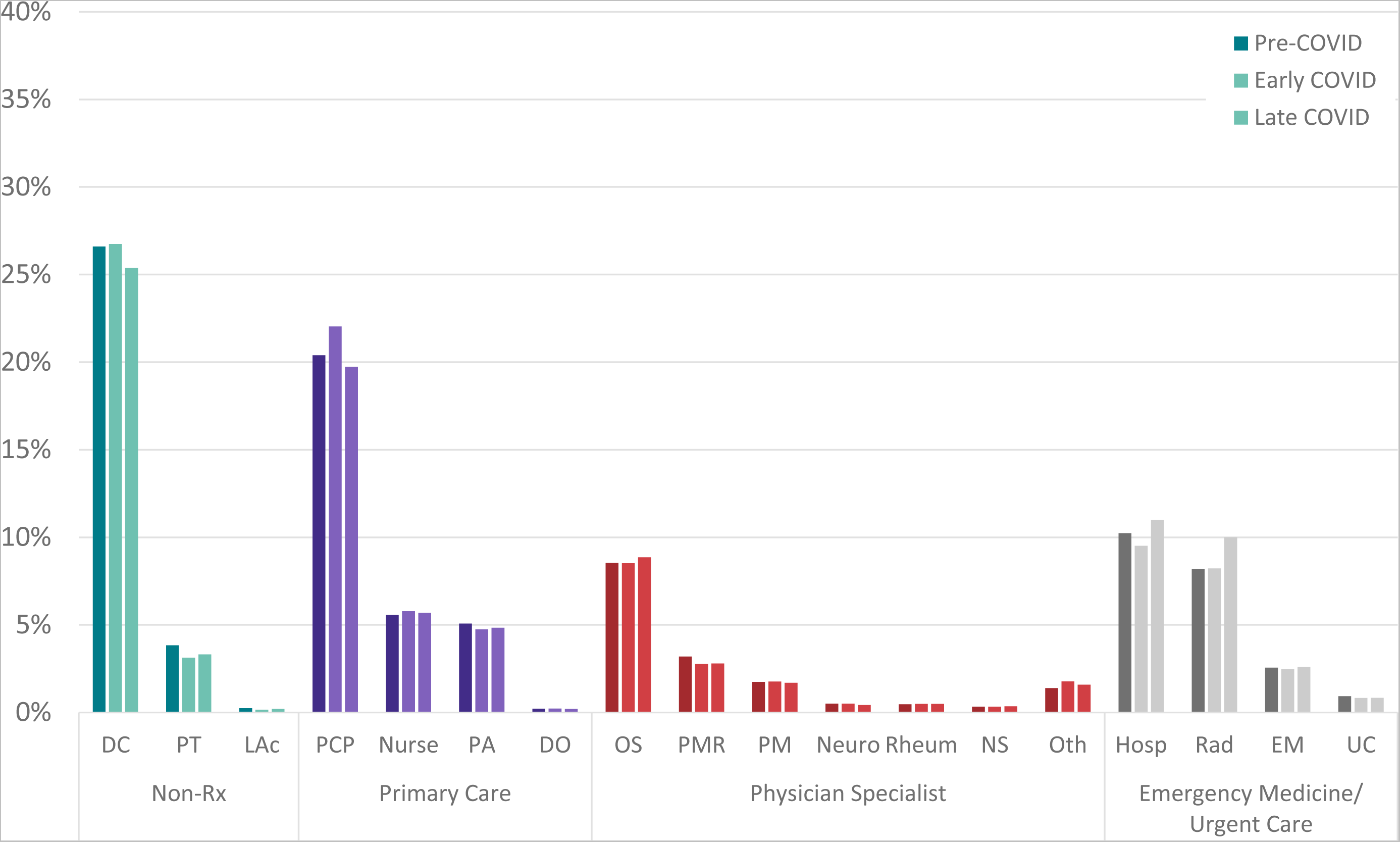
Distribution of episodes by type of healthcare provider initially contacted by **commercially insured** individuals with LBP with duration <91 days PCP=primary care provider, Nur=nurse, PA=physician assistant, DO=doctor of osteopathy, DC=doctor of chiropractic, PT=physical therapist, LAc=licensed acupuncturist, OS=orthopedic suregon, PMR=physical medicine and rehabilitation, PM=pain managment, Neuro=neurologist, Rheum=rheumatologist, NS=neurosurgeon, Oth=other physician specialist, Hosp=hospital, Rad=radiologist, EM=emergency medicine, UC=urgent care

**Figure 2.**
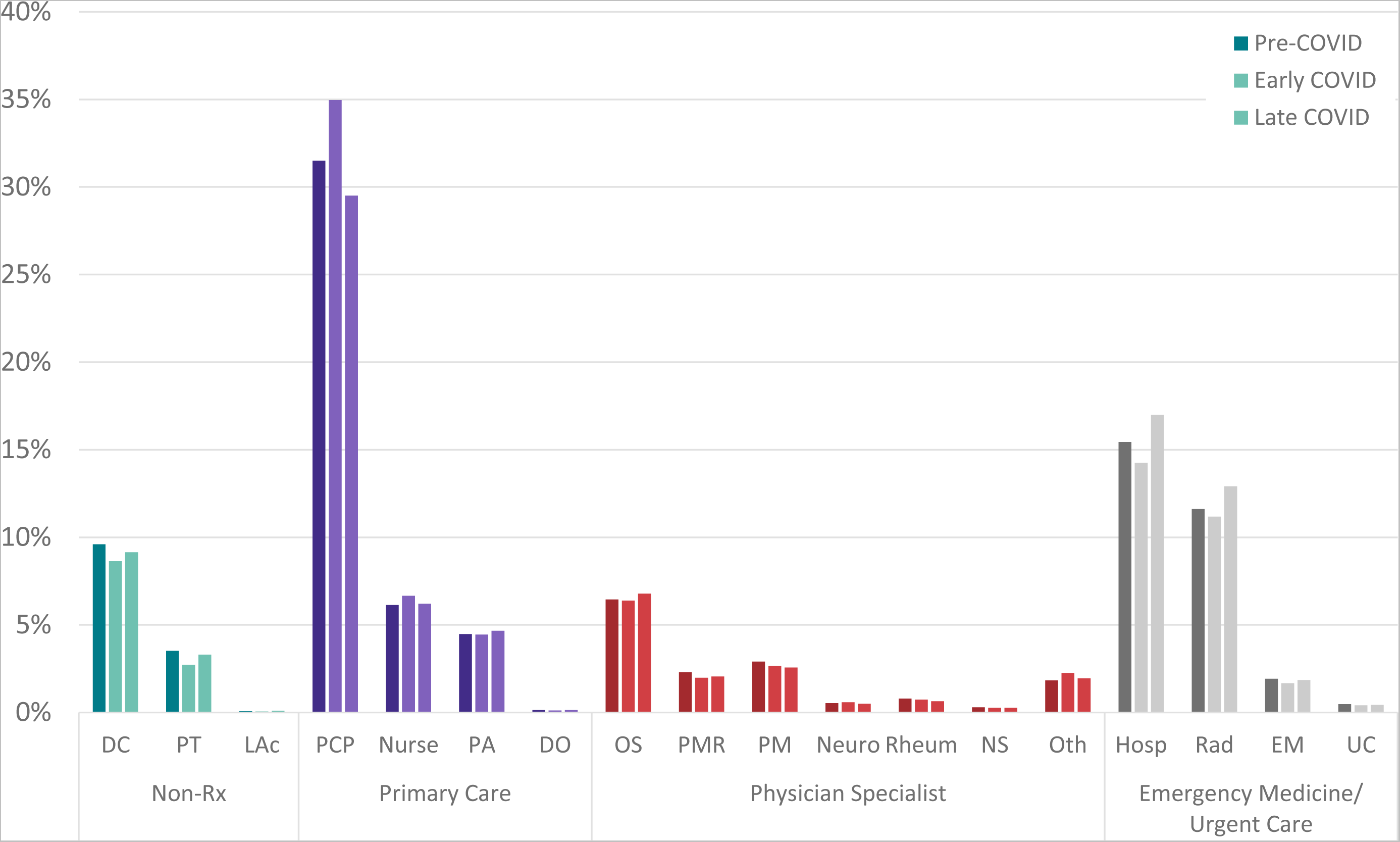
Distribution of episodes by type of healthcare provider initially contacted by **medicare advantage insured** individuals with LBP with duration <91 days PCP=primary care provider, Nur=nurse, PA=physician assistant, DO=doctor of osteopathy, DC=doctor of chiropractic, PT=physical therapist, LAc=licensed acupuncturist, OS=orthopedic suregon, PMR=physical medicine and rehabilitation, PM=pain managment, Neuro=neurologist, Rheum=rheumatologist, NS=neurosurgeon, Oth=other physician specialist, Hosp=hospital, Rad=radiologist, EM=emergency medicine, UC=urgent care

COVID was associated with significant changes in the type of HCP seen initially by individuals with LBP. In the CI cohort PTs, LAcs, and PMR physicians were initially contacted less often in the early and late COVID periods. In the late COVID period initial contact with hospitals and radiologists increased. [Figure 3] In the MA cohort, initial contact with DCs, PTs, PMR, PM, EM and UC were all lower in the early and late COVID periods. In the early COVID period initial contact with PCPs, nurses, and neurologists increased. In the late COVID period initial contact with LAcs, hospitals and radiologists increased. [Figure 4] [Table 2]

**Figure 3.**
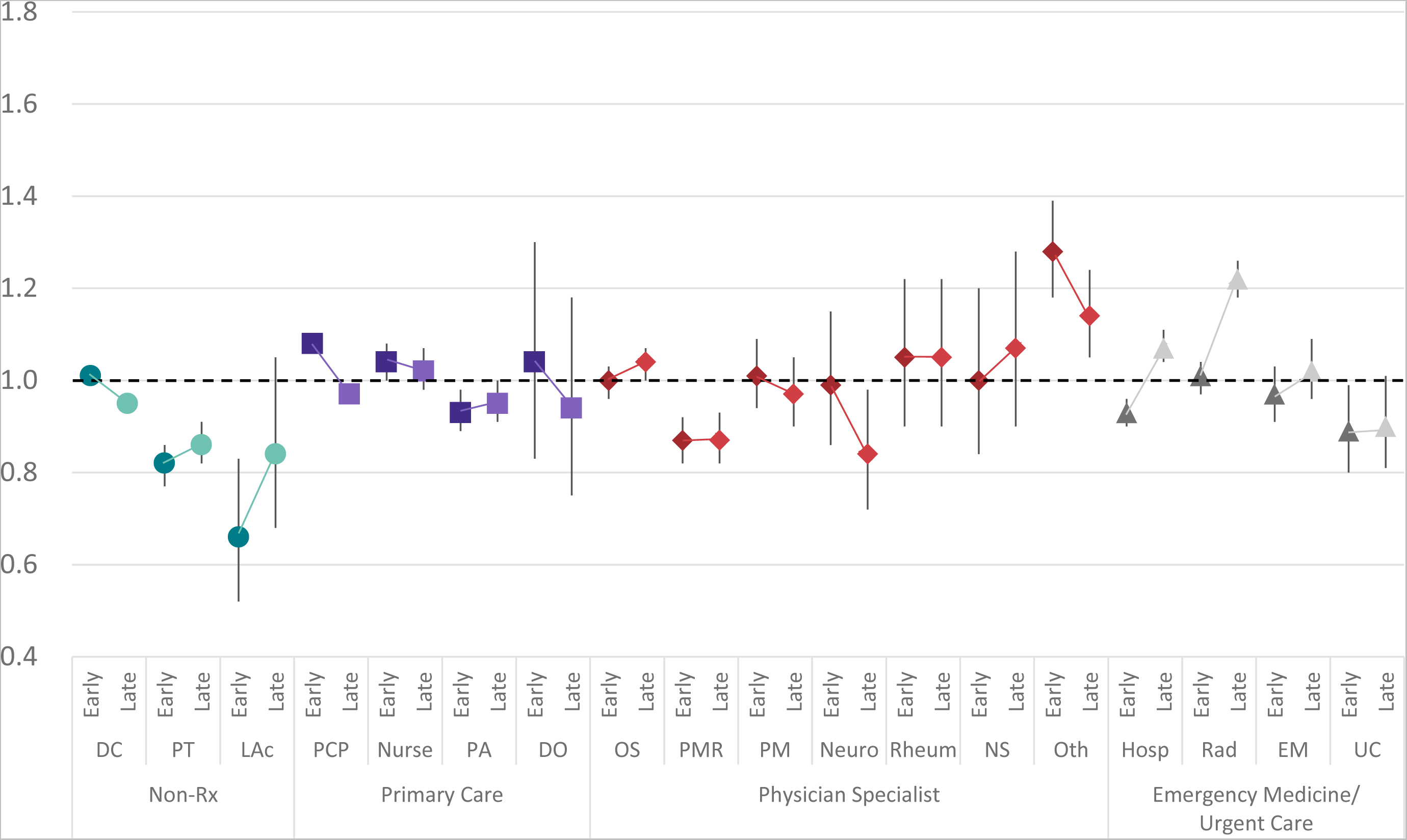
Risk ratio and 95% confidence interval comparing **early COVID and late-COVID** periods to pre-COVID period for type of health care provider initially contacted by **commercially insured** individuals with low back pain with episode lasting <91 days C=commercial insurance, M=medicare advantage, PCP=primary care provider, Nur=nurse, PA=physician assistant, DO=doctor of osteopathy, DC=doctor of chiropractic, PT=physical therapist, LAc=licensed acupuncturist, OS=orthopedic suregon, PMR=physical medicine and rehabilitation, PM=pain managment, Neuro=neurologist, Rheum=rheumatologist, NS=neurosurgeon, Oth=other physician specialist, Hosp=hospital, Rad=radiologist, EM=emergency medicine, UC=urgent care

**Figure 4.**
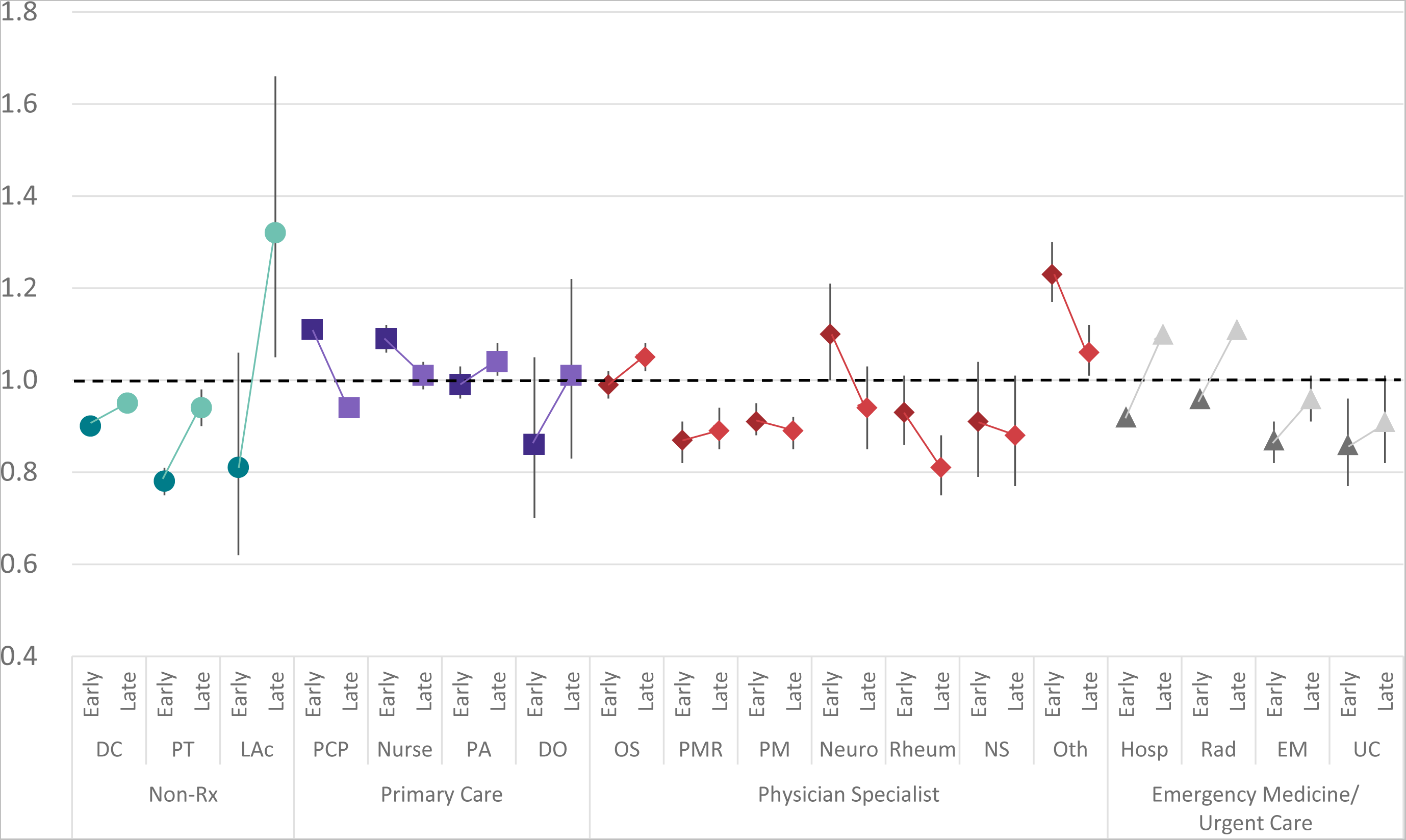
Risk ratio and 95% confidence interval comparing **early COVID and late-COVID** periods to pre-COVID period for type of health care provider initially contacted by **medicare advantage insured** individuals with low back pain with episode lasting <91 days C=commercial insurance, M=medicare advantage, PCP=primary care provider, Nur=nurse, PA=physician assistant, DO=doctor of osteopathy, DC=doctor of chiropractic, PT=physical therapist, LAc=licensed acupuncturist, OS=orthopedic suregon, PMR=physical medicine and rehabilitation, PM=pain managment, Neuro=neurologist, Rheum=rheumatologist, NS=neurosurgeon, Oth=other physician specialist, Hosp=hospital, Rad=radiologist, EM=emergency medicine, UC=urgent care

In the CI cohort, during the pre, early, and late COVID periods plain film radiology, chiropractic manipulative treatment (CMT), active care (AC), prescription NSAIDs, and prescription skeletal muscle relaxants were the only services provided for at least 15% of episodes. [Figure 5] In the MA cohort, during the pre, early, and late COVID periods plain film radiology, prescription NSAIDs, and prescription opioids were provided during at least 15% of episodes. In the late COVID period MRI was provided for 16.9% of episodes in the MA cohort. [Figure 6] [Table 3]

**Figure 5.**
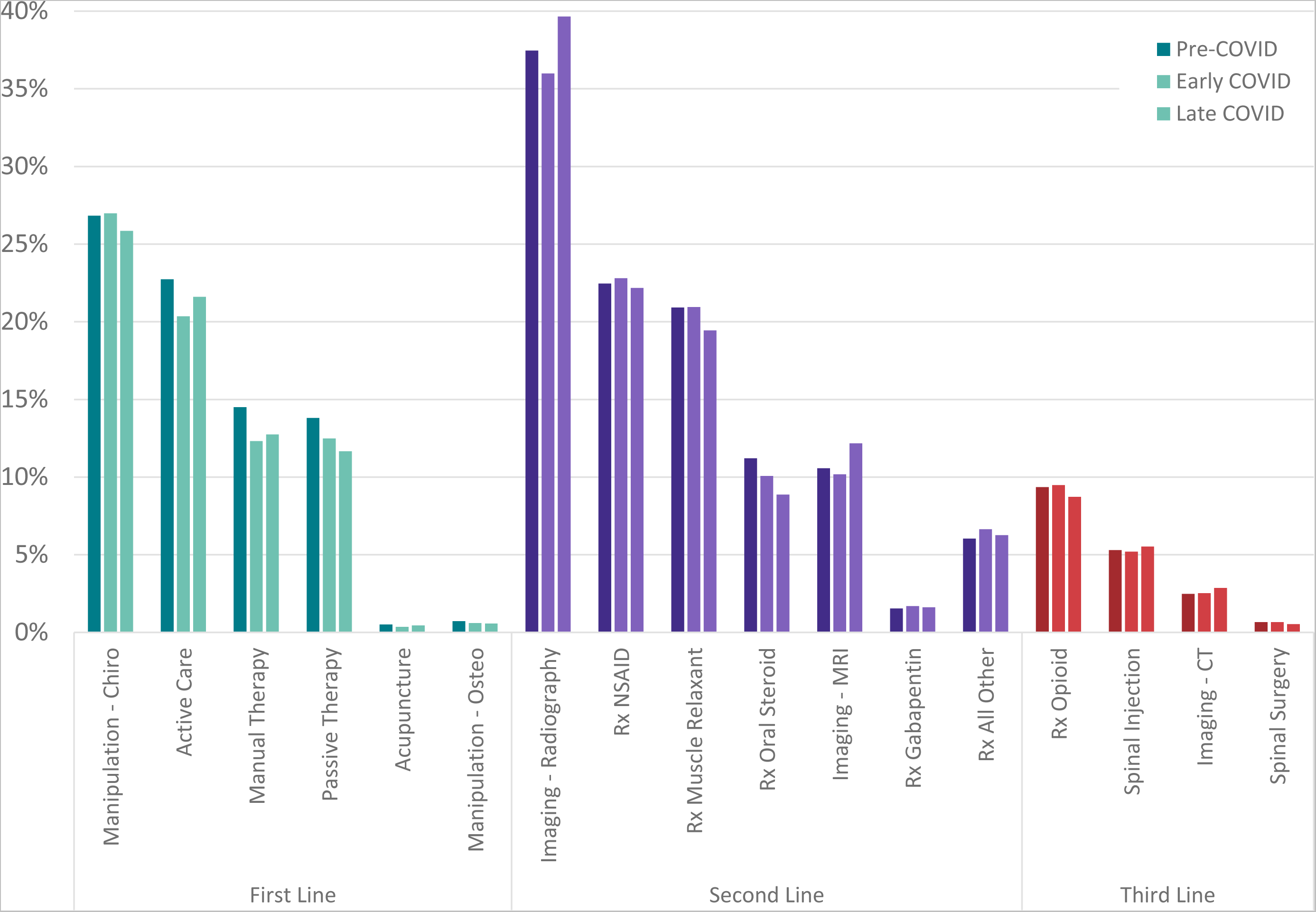
% of episodes including specific services provided for **commercially insured** individuals with low back pain with duration <91 days

**Figure 6.**
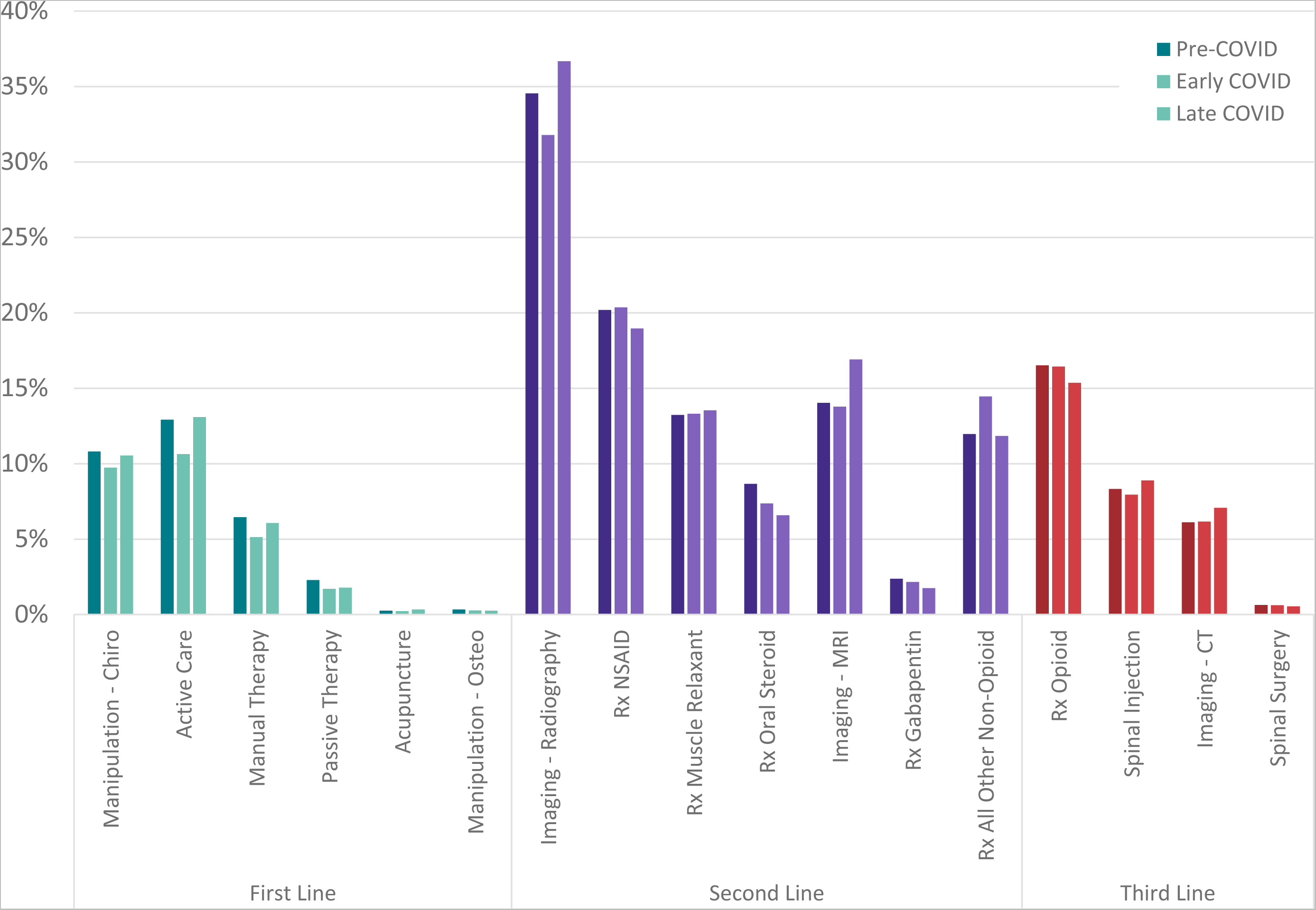
% of episodes including specific services provided for **medicare advantage** individuals with low back pain with duration <91 days

COVID was associated with significant changes in the services received by individuals with LBP. In the CI cohort, guideline concordant first line services like AC, manual therapy (MT), acupuncture, and osteopathic manipulative therapy were provided less often during COVID.

Prescription oral steroid were also provided less often during COVID. A significant increase in plain film radiology, MRI, and CT scans occurred in the late COVID period. [Figure 7] The same general pattern was observed in the MA cohort where guideline concordant first-line services decreased, particularly in the early COVID period. Reductions were also observed for prescription oral steroids and gabapentins in the early and late COVID periods. The percent of episodes with plain film radiography, MRI and CT scans, and spinal injections increased in the late COVID period. [Figure 8] [Table 3]

**Figure 7.**
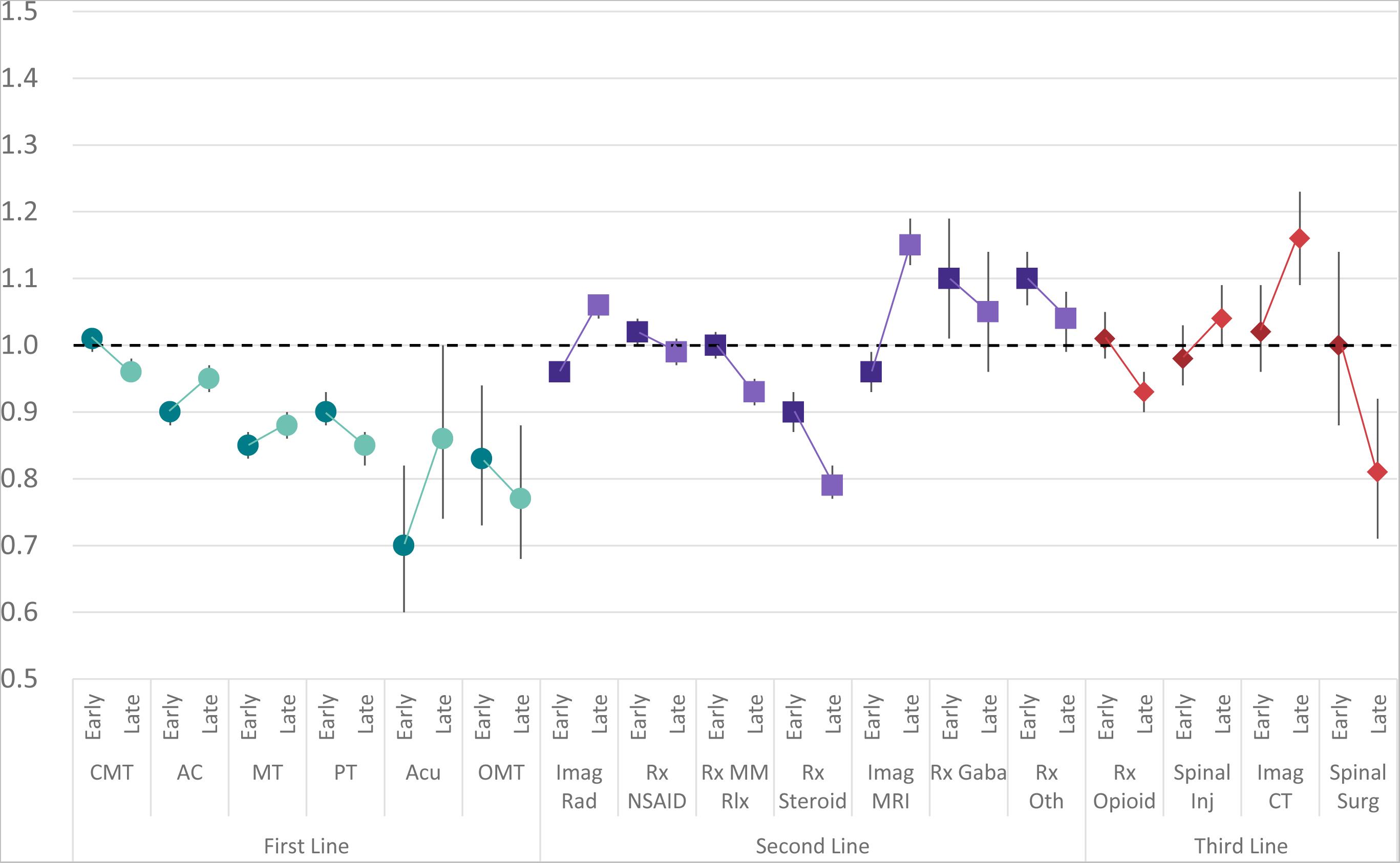
Risk ratio and 95% confidence interval comparing **early COVID and late COVID** period to pre-COVID period services provided for **commercially insured** individuals with low back pain with duration <91 days C=commercial insurance, M=medicare advantage, CMT=chiropractic manipulative therapy, AC=active care, MT=manual therapy, PT=passive therapy, Acu=acupuncture, OMT=osteopathic manipulative therapy, Imag Rad=radiology, MM Rlx=skeletal muscle relaxant, Imag MRI=MRI scan, Gaba=gabapentin, Oth=other prescription medication, Inj=injection, Imag-CT=CT scan, Surg=surgical procedure

**Figure 8.**
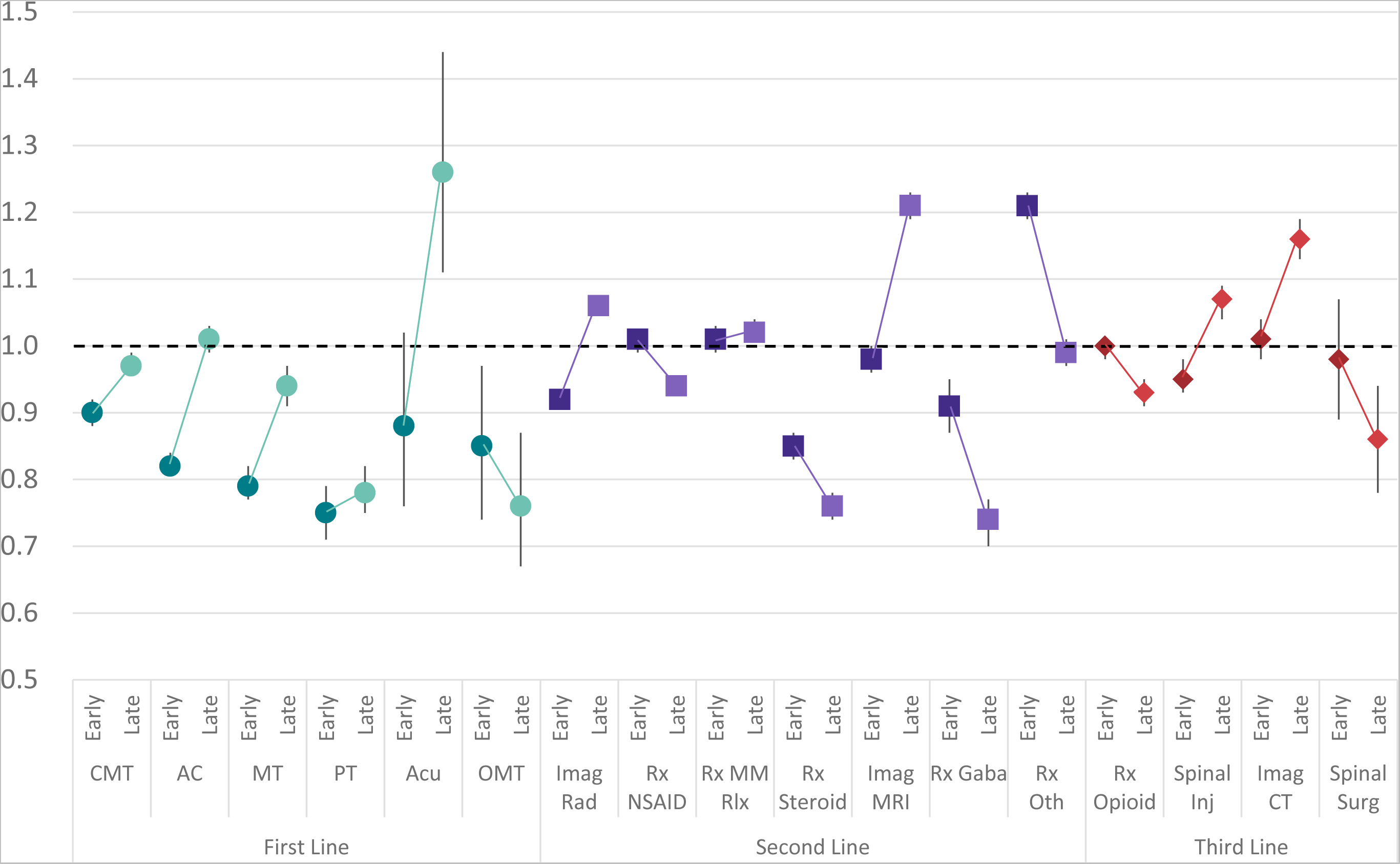
Risk ratio and 95% confidence interval comparing **early COVID and late COVID** period to pre-COVID period services provided for **medicare advantage insured** individuals with low back pain with duration <91 days C=commercial insurance, M=medicare advantage, CMT=chiropractic manipulative therapy, AC=active care, MT=manual therapy, PT=passive therapy, Acu=acupuncture, OMT=osteopathic manipulative therapy, Imag Rad=radiology, MM Rlx=skeletal muscle relaxant, Imag MRI=MRI scan, Gaba=gabapentin, Oth=other prescription medication, Inj=injection, Imag-CT=CT scan, Surg=surgical procedure

Differences between B25, Mixed and T25 cohorts in the Pre-, Early, and Late COVID period for both CI and MA were identified. In all time-periods in both CI and MA compared to the T25 cohort the B25 and Mixed cohorts were from zip codes with significantly higher ADI and significantly lower AGI. In all time-periods in both CI and MA compared to the T25 cohort the Mixed cohort was from zip codes with significantly higher percent non-Hispanic white (NHW) population. [Supplement – Table 1 – Stringency]

During the pre-COVID period and compared to the T25 reference, the type of HCP initially contacted by individuals with LBP was significantly different in the B25 and Mixed cohorts. [Figure 9] During the pre-COVID period compared to the T25 reference, the type of services provided to individuals with LBP was also significantly different in the B25 and Mixed cohorts. Most notably, first-line services were provided in a lower percent of episodes and third-line services per provided in a higher percent of episodes with variable differences in CI and MA. [Figure 10] [Supplement – Table 2 – Stringency] [Supplement – Table 3 – Stringency]

**Figure 9.**
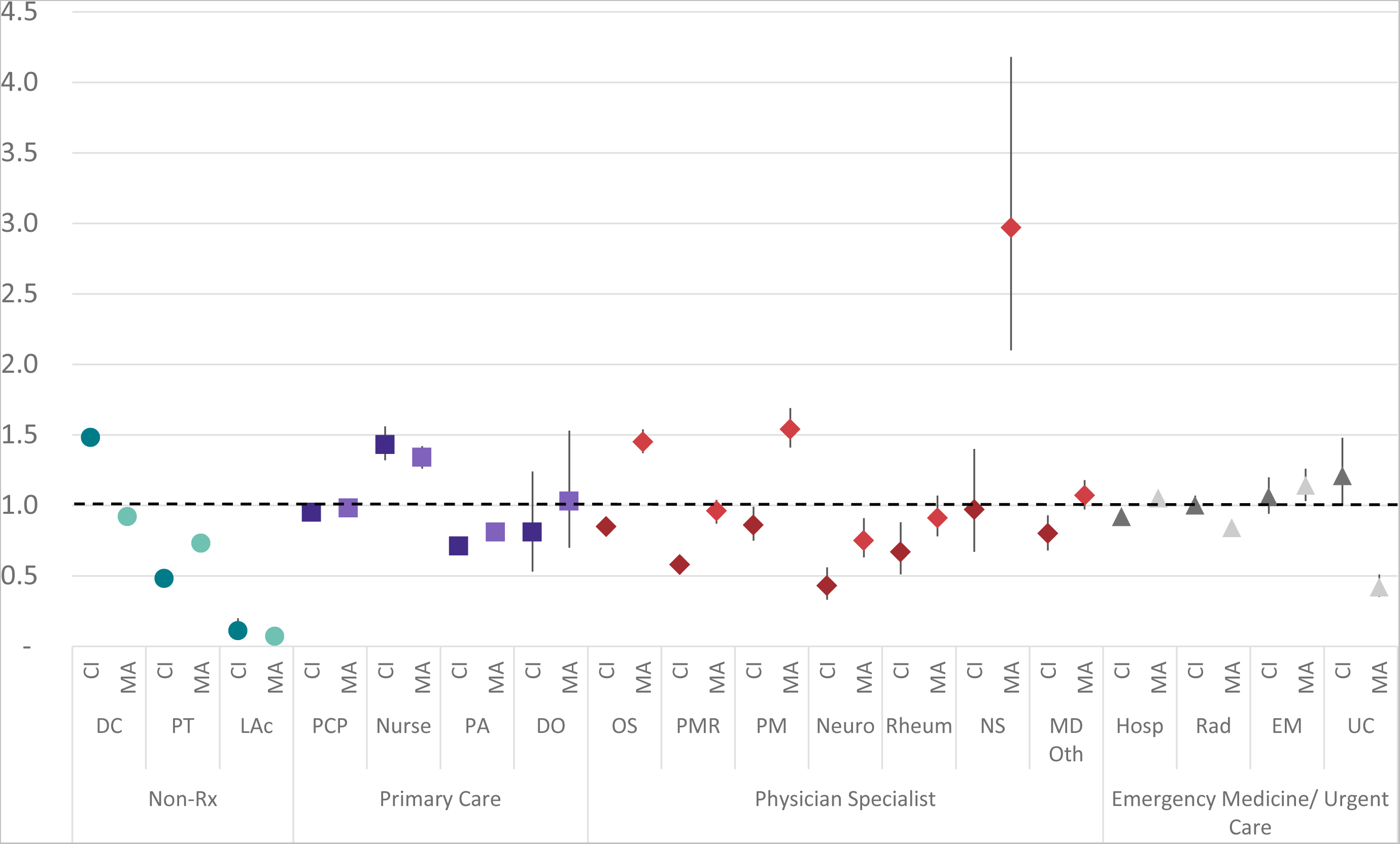
Risk ratio and 95% confidence interval comparing **B25 cohort to T25 cohort baseline** for the type of health care provider initially contacted by individuals with low back pain with episode lasting <91 days during the **pre-COVID period** B25=States in bottom 25 COVID public policy stringency response in early and late COVID period, T25=States in top 25, C=commercial insurance, M=medicare advantage, PCP=primary care provider, Nur=nurse, PA=physician assistant, DO=doctor of osteopathy, DC=doctor of chiropractic, PT=physical therapist, LAc=licensed acupuncturist, OS=orthopedic suregon, PMR=physical medicine and rehabilitation, PM=pain managment, Neuro=neurologist, Rheum=rheumatologist, NS=neurosurgeon, Oth=other physician specialist, Hosp=hospital, Rad=radiologist, EM=emergency medicine, UC=urgent care

**Figure 10.**
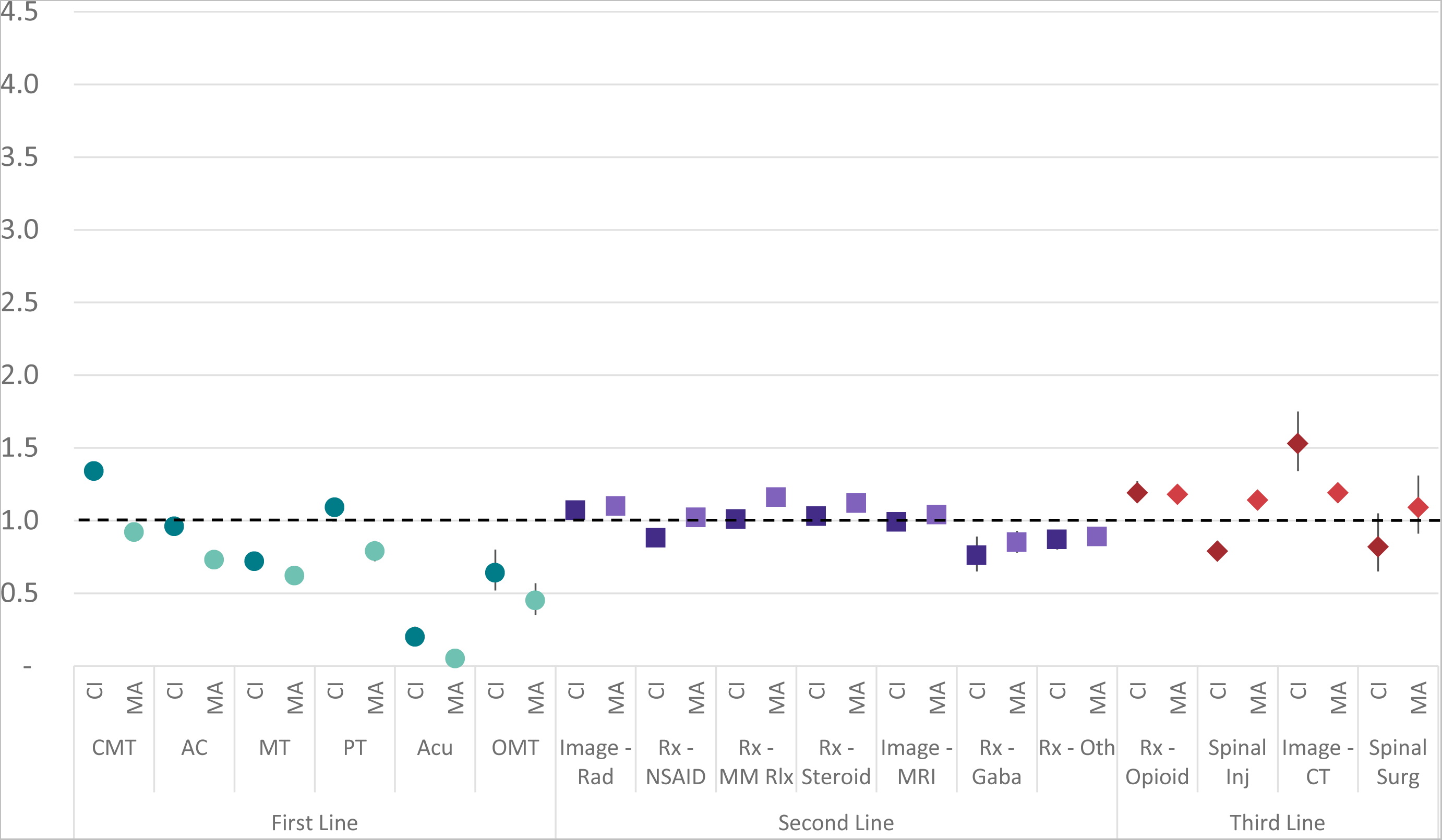
Risk ratio and 95% confidence interval comparing **B25 cohort to T25 cohort baseline** for the type of health care services provided for individuals with low back pain with episode lasting <91 days during the **pre-COVID period** B25=States in bottom 25 COVID public policy stringency response in early and late COVID period, T25=States in top 25, C=commercial insurance, M=medicare advantage, CMT=chiropractic manipulative therapy, AC=active care, MT=manual therapy, PT=passive therapy, Acu=acupuncture, OMT=osteopathic manipulative therapy, Imag Rad=radiology, MM Rlx=skeletal muscle relaxant, Imag MRI=MRI scan, Gaba=gabapentin, Oth=other prescription medication, Inj=injection, Imag-CT=CT scan, Surg=surgical procedure

There was no indication that variable COVID State public policy response stringency was associated with clinically significant differences in either the type of HCP contacted initially by individuals with LBP [Figure 11] or in the services provided for LBP. [Figure 12] A reduction in first-line services provided for LBP was observed in B25, Mixed and T25 cohorts. [Supplement – Table 2 – Stringency] [Supplement – Table 3 – Stringency]

**Figure 11.**
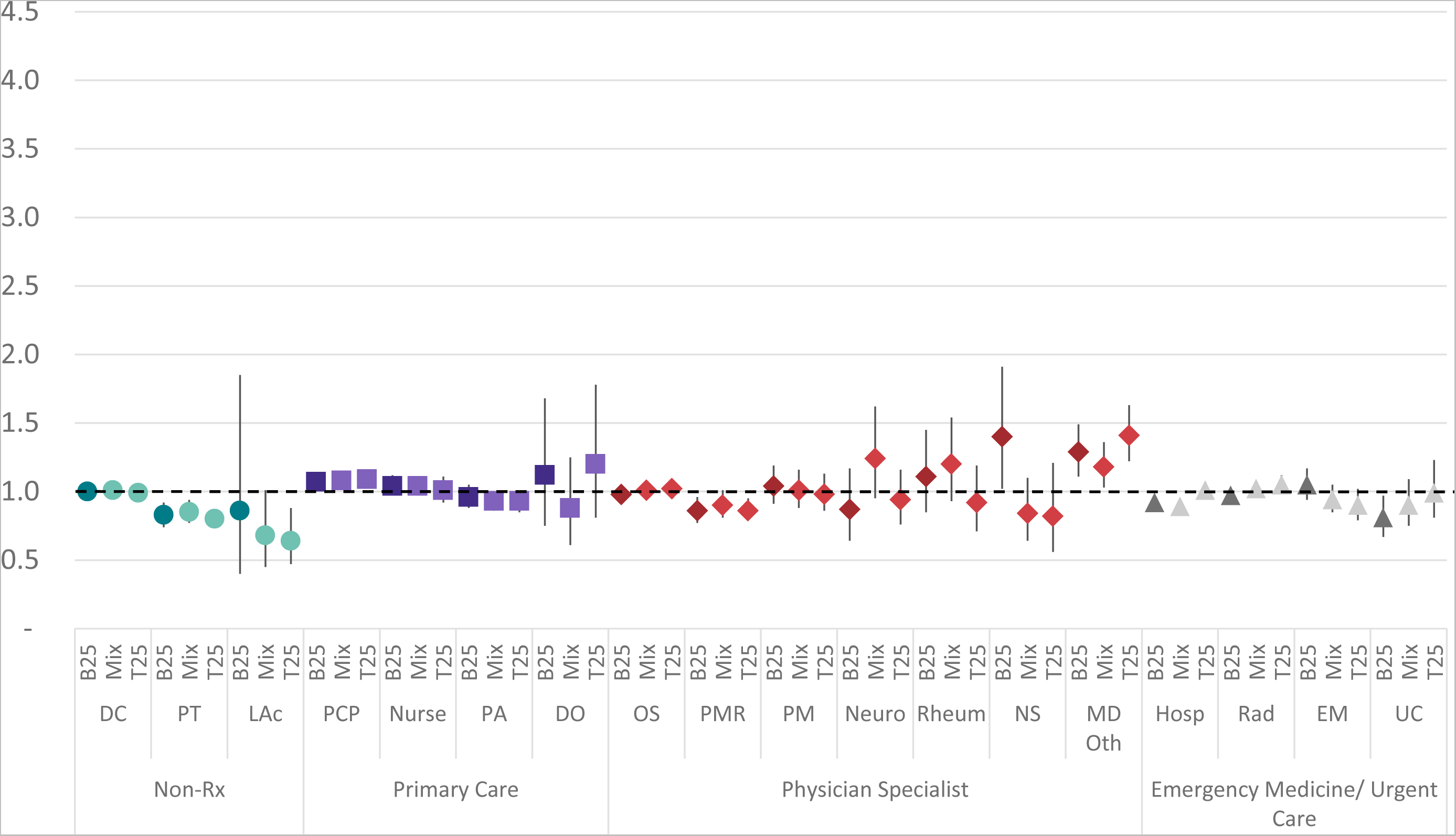
Risk ratio and 95% confidence interval comparing **the early COVID period to the pre-COVID baseline** for the type of health care provider initially contacted by **commercially insured** individuals with low back pain with episode lasting <91 days B25=States in bottom 25 COVID public policy stringency response in early and late COVID period, T25=States in top 25, Mix=States that alternated between B25 and T25 in early and late COVID periods, PCP=primary care provider, Nur=nurse, PA=physician assistant, DO=doctor of osteopathy, DC=doctor of chiropractic, PT=physical therapist, LAc=licensed acupuncturist, OS=orthopedic suregon, PMR=physical medicine and rehabilitation, PM=pain managment, Neuro=neurologist, Rheum=rheumatologist, NS=neurosurgeon, Oth=other physician specialist, Hosp=hospital, Rad=radiologist, EM=emergency medicine, UC=urgent care

**Figure 12.**
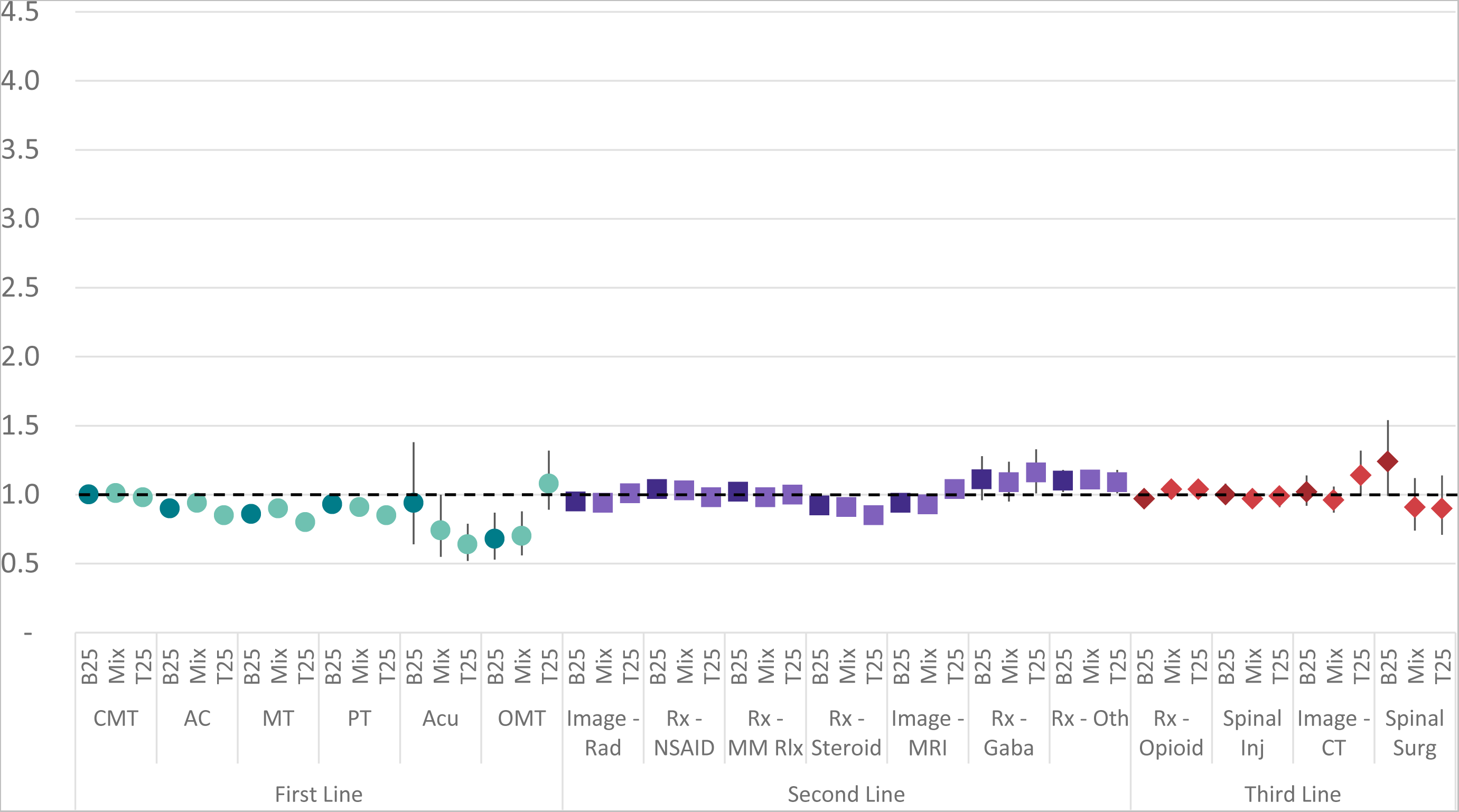
Risk ratio and 95% confidence interval comparing **the early COVID period to the pre-COVID baseline** for the type of health care services provided for **commercially insured** individuals with low back pain with episode lasting <91 days B25=States in bottom 25 COVID public policy stringency response in early and late COVID period, T25=States in top 25, Mix=States that alternated between B25 and T25 in early and late COVID periods, CMT=chiropractic manipulative therapy, AC=active care, MT=manual therapy, PT=passive therapy, Acu=acupuncture, OMT=osteopathic manipulative therapy, Imag Rad=radiology, MM Rlx=skeletal muscle relaxant, Imag MRI=MRI scan, Gaba=gabapentin, Oth=other prescription medication, Inj=injection, Imag-CT=CT scan, Surg=surgical procedure

## Discussion

The changes identified in care seeking for and management of LBP add to the growing understanding of the impact of the COVID pandemic. During COVID a greater proportion of CI and MA individuals with LBP initially contacted PCPs, nurses, and non-operative physician specialists while initial contact with DCs PT, OS, EM, and UC decreased. Early and late in the COVID pandemic fewer episodes included guideline concordant first-line services. In both the CI and MA cohorts use of low-value and potentially non-guideline concordant diagnostic imaging for LBP was higher in the late COVID period than before the onset of COVID. Variability in stringency of State public policy responses to COVID did not appear to have had an influence on observed changes. With the increase in total episode cost in the late COVID period the increased rate of imaging for LBP during COVID warrants additional study.

As is the case with any retrospective observational analysis of administrative data, there are numerous limitations and potential confounders to consider. The COVID pandemic added additional or amplified existing well known confounders and limitations for analyzing the type of HCP initially contacted by an individual with LBP.^19^ Variation in the rate of COVID infections by date and geography^56^ and corresponding local COVID public health policies ^5, 6, 57–59^, coupled with the analysis not being a representative sample of the U.S. introduced numerous potential confounders beyond the scope of the analysis. The impact of avoidance of elective spine surgery ^60^, individuals reluctance to seek in-person hands on care, reduced capacity of primary care ^61^ and emergency departments ^62^, and reluctance to use of public transportation ^63, 64^ are a few examples. Confounding these confounders was the novel finding that in the baseline pre-COVID period the types of HCP initially contacted by individuals with LBP, and services provided for LBP, were significantly different among the B25, Mixed and T25 States.

The potential confounders associated with the heterogenous nature of CI and MA coverage and cohorts, differing proportion of CI and MA episodes from each State, and variable State public policy responses to COVID made it impossible to interpret observed differences in the type of HCP initially contacted by individuals with LBP and subsequent use of health care services. The potentially novel finding of the significant difference in episode duration in the pre, early and late COVID periods observed between CI and MA cohorts and addressed through limiting the study to episodes with a duration of less than 91 days duration, warrants additional investigation.

This study corroborated and contrasted with the findings of other studies exploring the impact of COVID on the management of LBP. Among non-pharmaceutical HCP types the proportion of individuals with LBP initially contacting a DC was unchanged in the CI cohort and exhibited the smallest reduction in the MA, a finding aligned with how DCs may have adapted practice.^65^ The observed increase in the proportion of individuals with LBP contacting a PCP in the US conflicts with an observed reduction in PCP access in Germany.^66^ Within the MA cohort the finding of a reduction in individuals with LBP initially contacting EM during COVID is aligned with previously observed reductions in patients with non-urgent conditions like back pain presenting to emergency departments in the US and Finland.^30, 67^ The current study did not find a reduction in CI individuals with LBP presenting to EM.

Studies conducted in the US and Canada found COVID-related changes in pharmacological pain treatments, including increased opioid use, during the pandemic with a contributing factor being decreased availability and/or use of non-pharmaceutical therapy options.^26, 27^ Similarly, a study of high and low value outpatient services early in the COVID pandemic found an increase in the use of prescription opioids for neck and back pain.^25^ Additional studies from the US found that use of non-pharmaceutical decreased for LBP, while and use of prescription opioids increased or were unchanged.^35, 36^ An analysis of interventional procedures provided for chronic pain in a Medicare cohort for a significant reduction associated with the COVID pandemic.^29^ The current study corroborated previous study findings of COVID being associated with a reduction in use of non-pharmaceutical services for LBP, but did not find an increase in use of prescription opioids. Similarly, the current study found the rate of interventional procedures, like spinal injection or surgery, was unchanged or lower during COVID, except for an increased rate of spinal injection in the late COVID period for the MA cohort.

## Conclusions

The COVID pandemic was associated with changes in the types of HCP initially contacted by and subsequent services provided to individuals with LBP. Among COVID related changes to services provided for LBP, in both the CI and MA cohorts the most consistent findings were a reduction in the rate of guideline concordant first-line services, and an increase in the rate of diagnostic imaging during the late COVID period.

## Declarations

### Ethics approval and consent to participate

Because the data was de-identified or a Limited Data Set in compliance with the Health Insurance Portability and Accountability Act and customer requirements, Institutional Review Board approval or waiver of authorization was not required.

### Consent for publication

Not applicable

### Availability of data and materials

The data are proprietary and are not available for public use but, under certain conditions, may be made available to editors and their approved auditors under a data-use agreement to confirm the findings of the current study.

### Authors’ contributions

Study conception and design; **DE**. Data acquisition; **DE, MZ.** Data analysis and interpretation;

**DE, MZ.** Draft or revise manuscript; **DE, MZ.**

## Supporting information

Supplement - Episode Duration

Supplement - Public Policy

Supplement - State

Supplement - STROBE Checklist

Supplement - Table 1 - Full Cohort

Supplement - Table 1 - Stringency

Supplement - Table 2 - Full Cohort

Supplement - Table 2 - Stringency

Supplement - Table 3 - Full Cohort

Supplement - Table 3 - Stringency

## Data Availability

All data produced in the present work are contained in the manuscript

## Abbreviations

**List of Abbreviations**
LBP: Low back pain
CI: Commercial Insurance
MA: Medicare Advantage
US: United States
WHO: World Health Organization
CPG: Clinical practice guideline
HCP: Health care provider
ADI: Area Deprivation Index
AGI: Adjusted Gross Income
NHW: Non-Hispanic White
STROBE: Strengthening the Reporting of Observational Studies in Epidemiology
ETG*^®^*: Episode Treatment Group*^®^*
ERG*^®^*: Episode Risk Group*^®^*
SD: Standard deviation
IQR: Interquartile range
RR: Risk ratio
Q1: 1^st^ Quartile
Q3: 3^rd^ Quartile
T25: Top 25 State with most stringent COVID restrictions on 4/1/2020 and 4/1/2021
B25: Bottom 25 State with least stringent COVID restrictions on 4/1/2020 and 4/1/2021
PCP: Primary care provider
PA: Physician’s Assistant
DC: Doctor of Chiropractic
PT: Physical Therapist
LAc: Licensed Acupuncturist
OS: Orthopedic Surgeon
PM: Pain Management
EM: Emergency Medicine
UC: Urgent Care
CMT: Chiropractic manipulative treatment
OMT: Osteopathic manipulative treatment

