## Supplement - Episode Duration for "Impact of the COVID-19 pandemic on low back pain management in commercially insured and Medicare Advantage cohorts. A retrospective cohort study"

| Supplement - Episode Duration | | | |
|---|---|---|---|
| | Pre-COVID | Early COVID | Late COVID |
| Original Cohort | | | |
| Episodes - Count | | | |
| CI | 150761 | 158123 | 124958 |
| MA | 530583 | 493573 | 368842 |
| Episode Duration - Median (Q1, Q3) | | | |
| CI | 124 (18, 294) | 93 (12, 248) | 54 (4, 149) |
| MA | 248 (79, 460) | 204 (59, 315) | 101 (22, 199) |
| Episodes With Duration <91 days | | | |
| Episodes - Count | | | |
| CI | 67158 | 78050 | 76835 |
| MA | 141258 | 152406 | 172461 |
| Episode Duration - Median (Q1, Q3) | | | |
| CI | 12 (1, 43) | 11 (1, 43) | 13 (1, 44) |
| MA | 15 (1, 49) | 13 (1, 49) | 16 (1, 50) |
| Episodes With Duration <61 days | | | |
| Episodes - Count | | | |
| CI | 57889 | 66716 | 65586 |
| MA | 117095 | 124549 | 140328 |
| Episode Duration - Median (Q1, Q3) | | | |
| CI | 6 (1, 29) | 5 (1, 28) | 6 (1, 29) |
| MA | 6 (1, 30) | 2 (1, 29) | 6 (1, 31) |

CI=commercial insurance, MA=medicare advantage
