## Supplement - State for "Impact of the COVID-19 pandemic on low back pain management in commercially insured and Medicare Advantage cohorts. A retrospective cohort study"

Supplement State- Episode distribution by individual home address 5-digit zip code State and insurance coverage

| | Episode Count | | | | | | | | | | | | % of Total Episodes | | | | | | | | | | | |
|---|---|---|---|---|---|---|---|---|---|---|---|---|---|---|---|---|---|---|---|---|---|---|---|---|
| | Total | | | | Commercial | | | | Medicare Advantage | | | | Total | | | | Commercial | | | | Medicare Advantage | | | |
| | Total | Pre | Early | Late | Total | Pre | Early | Late | Total | Pre | Early | Late | Total | Pre | Early | Late | Total | Pre | Early | Late | Total | Pre | Early | Late |
| Total | 688168 | 208416 | 230456 | 249296 | 222043 | 67158 | 78050 | 76835 | 466125 | 141258 | 152406 | 172461 | 100% | 100% | 100% | 100% | 100% | 100% | 100% | 100% | 100% | 100% | 100% | 100% |
| FL | 70055 | 22045 | 23535 | 24475 | 19144 | 5793 | 6816 | 6535 | 50911 | 16252 | 16719 | 17940 | 10.2% | 10.6% | 10.2% | 9.8% | 8.6% | 8.6% | 8.7% | 8.5% | 10.9% | 11.5% | 11.0% | 10.4% |
| TX | 66761 | 20158 | 22504 | 24099 | 24166 | 7206 | 8320 | 8640 | 42595 | 12952 | 14184 | 15459 | 9.7% | 9.7% | 9.8% | 9.7% | 10.9% | 10.7% | 10.7% | 11.2% | 9.1% | 9.2% | 9.3% | 9.0% |
| NY | 44878 | 14494 | 14703 | 15681 | 23634 | 7784 | 7776 | 8074 | 21244 | 6710 | 6927 | 7607 | 6.5% | 7.0% | 6.4% | 6.3% | 10.6% | 11.6% | 10.0% | 10.5% | 4.6% | 4.8% | 4.5% | 4.4% |
| CA | 43115 | 13828 | 14597 | 14690 | 13984 | 4279 | 4857 | 4848 | 29131 | 9549 | 9740 | 9842 | 6.3% | 6.6% | 6.3% | 5.9% | 6.3% | 6.4% | 6.2% | 6.3% | 6.2% | 6.8% | 6.4% | 5.7% |
| GA | 34852 | 11083 | 11612 | 12157 | 5685 | 1590 | 2083 | 2012 | 29167 | 9493 | 9529 | 10145 | 5.1% | 5.3% | 5.0% | 4.9% | 2.6% | 2.4% | 2.7% | 2.6% | 6.3% | 6.7% | 6.3% | 5.9% |
| IL | 34556 | 10564 | 11380 | 12612 | 11484 | 3363 | 4137 | 3984 | 23072 | 7201 | 7243 | 8628 | 5.0% | 5.1% | 4.9% | 5.1% | 5.2% | 5.0% | 5.3% | 5.2% | 4.9% | 5.1% | 4.8% | 5.0% |
| NC | 27782 | 8000 | 9445 | 10337 | 7717 | 2292 | 2793 | 2632 | 20065 | 5708 | 6652 | 7705 | 4.0% | 3.8% | 4.1% | 4.1% | 3.5% | 3.4% | 3.6% | 3.4% | 4.3% | 4.0% | 4.4% | 4.5% |
| WI | 27616 | 8520 | 9300 | 9796 | 6710 | 1997 | 2371 | 2342 | 20906 | 6523 | 6929 | 7454 | 4.0% | 4.1% | 4.0% | 3.9% | 3.0% | 3.0% | 3.0% | 3.0% | 4.5% | 4.6% | 4.5% | 4.3% |
| MO | 25637 | 7606 | 8628 | 9403 | 8949 | 2724 | 3222 | 3003 | 16688 | 4882 | 5406 | 6400 | 3.7% | 3.6% | 3.7% | 3.8% | 4.0% | 4.1% | 4.1% | 3.9% | 3.6% | 3.5% | 3.5% | 3.7% |
| AZ | 23297 | 7614 | 7611 | 8072 | 6007 | 1953 | 2110 | 1944 | 17290 | 5661 | 5501 | 6128 | 3.4% | 3.7% | 3.3% | 3.2% | 2.7% | 2.9% | 2.7% | 2.5% | 3.7% | 4.0% | 3.6% | 3.6% |
| OH | 20342 | 5909 | 6983 | 7450 | 7134 | 2126 | 2597 | 2411 | 13208 | 3783 | 4386 | 5039 | 3.0% | 2.8% | 3.0% | 3.0% | 3.2% | 3.2% | 3.3% | 3.1% | 2.8% | 2.7% | 2.9% | 2.9% |
| CT | 20080 | 6229 | 6407 | 7444 | 4916 | 1510 | 1695 | 1711 | 15164 | 4719 | 4712 | 5733 | 2.9% | 3.0% | 2.8% | 3.0% | 2.2% | 2.2% | 2.2% | 2.2% | 3.3% | 3.3% | 3.1% | 3.3% |
| NJ | 19488 | 6230 | 6197 | 7061 | 9024 | 2961 | 2939 | 3124 | 10464 | 3269 | 3258 | 3937 | 2.8% | 3.0% | 2.7% | 2.8% | 4.1% | 4.4% | 3.8% | 4.1% | 2.2% | 2.3% | 2.1% | 2.3% |
| IN | 19039 | 5394 | 6429 | 7216 | 4198 | 1258 | 1535 | 1405 | 14841 | 4136 | 4894 | 5811 | 2.8% | 2.6% | 2.8% | 2.9% | 1.9% | 1.9% | 2.0% | 1.8% | 3.2% | 2.9% | 3.2% | 3.4% |
| CO | 17613 | 5450 | 6048 | 6115 | 6267 | 1920 | 2209 | 2138 | 11346 | 3530 | 3839 | 3977 | 2.6% | 2.6% | 2.6% | 2.5% | 2.8% | 2.9% | 2.8% | 2.8% | 2.4% | 2.5% | 2.5% | 2.3% |
| SC | 14618 | 4463 | 5014 | 5141 | 1643 | 428 | 613 | 602 | 12975 | 4035 | 4401 | 4539 | 2.1% | 2.1% | 2.2% | 2.1% | 0.7% | 0.6% | 0.8% | 0.8% | 2.8% | 2.9% | 2.9% | 2.6% |
| WA | 13552 | 4277 | 4511 | 4764 | 2407 | 684 | 866 | 857 | 11145 | 3593 | 3645 | 3907 | 2.0% | 2.1% | 2.0% | 1.9% | 1.1% | 1.0% | 1.1% | 1.1% | 2.4% | 2.5% | 2.4% | 2.3% |
| AL | 13091 | 3441 | 4584 | 5066 | 695 | 194 | 253 | 248 | 12396 | 3247 | 4331 | 4818 | 1.9% | 1.7% | 2.0% | 2.0% | 0.3% | 0.3% | 0.3% | 0.3% | 2.7% | 2.3% | 2.8% | 2.8% |
| VA | 13052 | 3717 | 4417 | 4918 | 6616 | 2052 | 2324 | 2240 | 6436 | 1665 | 2093 | 2678 | 1.9% | 1.8% | 1.9% | 2.0% | 3.0% | 3.1% | 3.0% | 2.9% | 1.4% | 1.2% | 1.4% | 1.6% |
| TN | 11984 | 3400 | 4118 | 4466 | 4829 | 1323 | 1708 | 1798 | 7155 | 2077 | 2410 | 2668 | 1.7% | 1.6% | 1.8% | 1.8% | 2.2% | 2.0% | 2.2% | 2.3% | 1.5% | 1.5% | 1.6% | 1.5% |
| OR | 11178 | 3339 | 3805 | 4034 | 1899 | 562 | 724 | 613 | 9279 | 2777 | 3081 | 3421 | 1.6% | 1.6% | 1.7% | 1.6% | 0.9% | 0.8% | 0.9% | 0.8% | 2.0% | 2.0% | 2.0% | 2.0% |
| UT | 10536 | 3210 | 3483 | 3843 | 1607 | 423 | 581 | 603 | 8929 | 2787 | 2902 | 3240 | 1.5% | 1.5% | 1.5% | 1.5% | 0.7% | 0.6% | 0.7% | 0.8% | 1.9% | 2.0% | 1.9% | 1.9% |
| PA | 8877 | 2471 | 2972 | 3434 | 3963 | 1113 | 1386 | 1464 | 4914 | 1358 | 1586 | 1970 | 1.3% | 1.2% | 1.3% | 1.4% | 1.8% | 1.7% | 1.8% | 1.9% | 1.1% | 1.0% | 1.0% | 1.1% |
| AR | 8688 | 2575 | 2900 | 3213 | 2583 | 737 | 903 | 943 | 6105 | 1838 | 1997 | 2270 | 1.3% | 1.2% | 1.3% | 1.3% | 1.2% | 1.1% | 1.2% | 1.2% | 1.3% | 1.3% | 1.3% | 1.3% |
| IA | 8655 | 2471 | 3031 | 3153 | 4088 | 1219 | 1552 | 1317 | 4567 | 1252 | 1479 | 1836 | 1.3% | 1.2% | 1.3% | 1.3% | 1.8% | 1.8% | 2.0% | 1.7% | 1.0% | 0.9% | 1.0% | 1.1% |
| OK | 8457 | 2245 | 2808 | 3404 | 2379 | 715 | 866 | 798 | 6078 | 1530 | 1942 | 2606 | 1.2% | 1.1% | 1.2% | 1.4% | 1.1% | 1.1% | 1.1% | 1.0% | 1.3% | 1.1% | 1.3% | 1.5% |
| MD | 7743 | 2440 | 2590 | 2713 | 6964 | 2241 | 2372 | 2351 | 779 | 199 | 218 | 362 | 1.1% | 1.2% | 1.1% | 1.1% | 3.1% | 3.3% | 3.0% | 3.1% | 0.2% | 0.1% | 0.1% | 0.2% |
| MA | 6072 | 1617 | 1924 | 2531 | 1635 | 404 | 550 | 681 | 4437 | 1213 | 1374 | 1850 | 0.9% | 0.8% | 0.8% | 1.0% | 0.7% | 0.6% | 0.7% | 0.9% | 1.0% | 0.9% | 0.9% | 1.1% |
| NE | 5261 | 1523 | 1776 | 1962 | 1859 | 568 | 698 | 593 | 3402 | 955 | 1078 | 1369 | 0.8% | 0.7% | 0.8% | 0.8% | 0.8% | 0.8% | 0.9% | 0.8% | 0.7% | 0.7% | 0.7% | 0.8% |
| LA | 4805 | 1441 | 1682 | 1682 | 3787 | 1146 | 1352 | 1289 | 1018 | 295 | 330 | 393 | 0.7% | 0.7% | 0.7% | 0.7% | 1.7% | 1.7% | 1.7% | 1.7% | 0.2% | 0.2% | 0.2% | 0.2% |
| NM | 4424 | 1285 | 1496 | 1643 | 672 | 184 | 249 | 239 | 3752 | 1101 | 1247 | 1404 | 0.6% | 0.6% | 0.6% | 0.7% | 0.3% | 0.3% | 0.3% | 0.3% | 0.8% | 0.8% | 0.8% | 0.8% |
| ID | 4240 | 1196 | 1413 | 1631 | 251 | 54 | 84 | 113 | 3989 | 1142 | 1329 | 1518 | 0.6% | 0.6% | 0.6% | 0.7% | 0.1% | 0.1% | 0.1% | 0.1% | 0.9% | 0.8% | 0.9% | 0.9% |
| MN | 3426 | 743 | 1038 | 1645 | 1346 | 378 | 453 | 515 | 2080 | 365 | 585 | 1130 | 0.5% | 0.4% | 0.5% | 0.7% | 0.6% | 0.6% | 0.6% | 0.7% | 0.4% | 0.3% | 0.4% | 0.7% |
| KS | 3299 | 798 | 1064 | 1437 | 1297 | 388 | 496 | 413 | 2002 | 410 | 568 | 1024 | 0.5% | 0.4% | 0.5% | 0.6% | 0.6% | 0.6% | 0.6% | 0.5% | 0.4% | 0.3% | 0.4% | 0.6% |
| RI | 3035 | 969 | 961 | 1105 | 709 | 215 | 235 | 259 | 2326 | 754 | 726 | 846 | 0.4% | 0.5% | 0.4% | 0.4% | 0.3% | 0.3% | 0.3% | 0.3% | 0.5% | 0.5% | 0.5% | 0.5% |
| NV | 2997 | 676 | 1071 | 1250 | 1307 | 372 | 481 | 454 | 1690 | 304 | 590 | 796 | 0.4% | 0.3% | 0.5% | 0.5% | 0.6% | 0.6% | 0.6% | 0.6% | 0.4% | 0.2% | 0.4% | 0.5% |
| MI | 2886 | 869 | 1023 | 994 | 2199 | 645 | 822 | 732 | 687 | 224 | 201 | 262 | 0.4% | 0.4% | 0.4% | 0.4% | 1.0% | 1.0% | 1.1% | 1.0% | 0.1% | 0.2% | 0.1% | 0.2% |
| ME | 2812 | 837 | 917 | 1058 | 267 | 75 | 101 | 91 | 2545 | 762 | 816 | 967 | 0.4% | 0.4% | 0.4% | 0.4% | 0.1% | 0.1% | 0.1% | 0.1% | 0.5% | 0.5% | 0.5% | 0.6% |
| KY | 2752 | 758 | 969 | 1025 | 1645 | 445 | 608 | 592 | 1107 | 313 | 361 | 433 | 0.4% | 0.4% | 0.4% | 0.4% | 0.7% | 0.7% | 0.8% | 0.8% | 0.2% | 0.2% | 0.2% | 0.3% |
| NH | 2655 | 707 | 863 | 1085 | 345 | 76 | 123 | 146 | 2310 | 631 | 740 | 939 | 0.4% | 0.3% | 0.4% | 0.4% | 0.2% | 0.1% | 0.2% | 0.2% | 0.5% | 0.4% | 0.5% | 0.5% |
| MS | 2258 | 598 | 803 | 857 | 1861 | 519 | 709 | 633 | 397 | 79 | 94 | 224 | 0.3% | 0.3% | 0.3% | 0.3% | 0.8% | 0.8% | 0.9% | 0.8% | 0.1% | 0.1% | 0.1% | 0.1% |
| WV | 1475 | 332 | 495 | 648 | 522 | 153 | 188 | 181 | 953 | 179 | 307 | 467 | 0.2% | 0.2% | 0.2% | 0.3% | 0.2% | 0.2% | 0.2% | 0.2% | 0.2% | 0.1% | 0.2% | 0.3% |
| VT | 1401 | 397 | 468 | 536 | 49 | 11 | 16 | 22 | 1352 | 386 | 452 | 514 | 0.2% | 0.2% | 0.2% | 0.2% | 0.0% | 0.0% | 0.0% | 0.0% | 0.3% | 0.3% | 0.3% | 0.3% |
| HI | 1086 | 323 | 359 | 404 | 56 | 16 | 23 | 17 | 1030 | 307 | 336 | 387 | 0.2% | 0.2% | 0.2% | 0.2% | 0.0% | 0.0% | 0.0% | 0.0% | 0.2% | 0.2% | 0.2% | 0.2% |
| DC | 790 | 279 | 243 | 268 | 752 | 265 | 232 | 255 | 38 | 14 | 11 | 13 | 0.1% | 0.1% | 0.1% | 0.1% | 0.3% | 0.4% | 0.3% | 0.3% | 0.0% | 0.0% | 0.0% | 0.0% |
| DE | 600 | 119 | 181 | 300 | 292 | 71 | 122 | 99 | 308 | 48 | 59 | 201 | 0.1% | 0.1% | 0.1% | 0.1% | 0.1% | 0.1% | 0.2% | 0.1% | 0.1% | 0.0% | 0.0% | 0.1% |
| SD | 451 | 76 | 142 | 233 | 154 | 41 | 51 | 62 | 297 | 35 | 91 | 171 | 0.1% | 0.0% | 0.1% | 0.1% | 0.1% | 0.1% | 0.1% | 0.1% | 0.1% | 0.0% | 0.1% | 0.1% |
| VI | 359 | 71 | 113 | 175 | 224 | 70 | 85 | 69 | 135 | 1 | 28 | 106 | 0.1% | 0.0% | 0.0% | 0.1% | 0.1% | 0.1% | 0.1% | 0.1% | 0.0% | 0.0% | 0.0% | 0.1% |
| WY | 355 | 112 | 127 | 116 | 156 | 52 | 60 | 44 | 199 | 60 | 67 | 72 | 0.1% | 0.1% | 0.1% | 0.0% | 0.1% | 0.1% | 0.1% | 0.1% | 0.0% | 0.0% | 0.0% | 0.0% |
| MT | 302 | 74 | 95 | 133 | 117 | 26 | 43 | 48 | 185 | 48 | 52 | 85 | 0.0% | 0.0% | 0.0% | 0.1% | 0.1% | 0.0% | 0.1% | 0.1% | 0.0% | 0.0% | 0.0% | 0.0% |
| ND | 253 | 44 | 95 | 114 | 137 | 27 | 56 | 54 | 116 | 17 | 39 | 60 | 0.0% | 0.0% | 0.0% | 0.0% | 0.1% | 0.0% | 0.1% | 0.1% | 0.0% | 0.0% | 0.0% | 0.0% |
| AK | 65 | 25 | 24 | 16 | 41 | 18 | 15 | 8 | 24 | 7 | 9 | 8 | 0.0% | 0.0% | 0.0% | 0.0% | 0.0% | 0.0% | 0.0% | 0.0% | 0.0% | 0.0% | 0.0% | 0.0% |
| PR | 62 | 12 | 27 | 23 | 44 | 6 | 20 | 18 | 18 | 6 | 7 | 5 | 0.0% | 0.0% | 0.0% | 0.0% | 0.0% | 0.0% | 0.0% | 0.0% | 0.0% | 0.0% | 0.0% | 0.0% |
| Unknown | 4505 | 1362 | 1475 | 1668 | 1627 | 486 | 570 | 571 | 2878 | 876 | 905 | 1097 | 0.7% | 0.7% | 0.6% | 0.7% | 0.7% | 0.7% | 0.7% | 0.7% | 0.6% | 0.6% | 0.6% | 0.6% |
