## Supplement - Table 1 - Full Cohort for "Impact of the COVID-19 pandemic on low back pain management in commercially insured and Medicare Advantage cohorts. A retrospective cohort study"

| Supplement - Table 1 - Cohort and total episode attributes - Full Cohort | | | |
|---|---|---|---|
| Actuals or Median (Q1, Q3) | Pre-COVID | Early COVID | Late COVID |
| Commercial Insurance (CI) | | | |
| Episodes | 150761 | 158123 | 124958 |
| # Unique HCPs | 87869 | 92951 | 78634 |
| Total Cost | 612354099 | 499764636 | 247289922 |
| Individuals | 141338 | 149204 | 124390 |
| Individuals - % Female | 54.0% | 53.3% | 53.2% |
| Individuals - Age | 48 (36, 57) | 48 (36, 57) | 48 (35, 57) |
| Individuals - ERG® Risk Score | 1.6 (0.7, 3.3) | 1.6 (0.7, 3.3) | 1.6 (0.7, 3.2) |
| Population - % NHW | 66.7% (48.1%, 79.6%) | 66.8% (47.9%, 79.9%) | 66.4% (46.8%, 79.6%) |
| Population - ADI | 40 (21, 60) | 41 (22, 61) | 40 (21, 60) |
| Population - AGI (1000s) | 74 (55, 110) | 73 (55, 107) | 74 (55, 110) |
| Episode - Total Cost | $560 (170, 1937) | $447 (134, 1536) | $376 (120, 1168) |
| Episode - # of Different HCP Seen | 2 (1, 3) | 2 (1, 3) | 2 (1, 3) |
| Episode - Duration | 124 (18, 294) | 93 (12, 248) | 54 (4, 149) |
| Medicare Advantage (MA) | | | |
| Episodes | 530583 | 493573 | 368842 |
| # Unique HCPs | 167568 | 167963 | 148372 |
| Total Cost | 1552372793 | 992871562 | 438435559 |
| Individuals | 491401 | 455171 | 368022 |
| Individuals - % Female | 62.1% | 61.4% | 61.3% |
| Individuals - Age | 72 (66, 77) | 72 (67, 78) | 73 (68, 78) |
| Individuals - ERG® Risk Score | 1.1 (0.4, 1.9) | 1.1 (0.5, 2.0) | 1.2 (0.6, 2.0) |
| Population - % NHW | 67.3% (46.1%, 81.3%) | 68.4% (47.0%, 81.8%) | 68.9% (47.5%, 81.9%) |
| Population - ADI | 55 (36, 72) | 55 (36, 72) | 54 (35, 71) |
| Population - AGI (1000s) | 59 (48, 79) | 60 (49, 79) | 61 (49, 82) |
| Episode - Total Cost | $579 (141, 1983) | $398 (104, 1351) | $281 (75, 891) |
| Episode - # of Different HCP Seen | 2 (1, 4) | 2 (1, 3) | 2 (1, 3) |
| Episode - Duration | 248 (79, 460) | 204 (59, 315) | 101 (22, 199) |

Within CI and MA, compared to Pre-COVID baseline, cells in red are not significantly different - Mann-Whitney U test (p=0.05)

Comparing MA to CI, all MA measures were signficantly different from CI measures - Mann-Whitney U test (p=0.05)
