## Supplement - Table 1 - Stringency for "Impact of the COVID-19 pandemic on low back pain management in commercially insured and Medicare Advantage cohorts. A retrospective cohort study"

### Supplement - Table 1 - Cohort Attributes By Pre-, Early and Late COVID Periods and Stringency of State Public Policy Response

| | Pre-COVID | | | Early COVID | | | Late COVID | | |
|---|---|---|---|---|---|---|---|---|---|
| (Median (IQR) (Q1,Q3)) | Bottom 25 | Mixed | Top 25 - reference | Bottom 25 | Mixed | Top 25 - reference | Bottom 25 | Mixed | Top 25 - reference |
| | Commercial Insurance (CI) | | | | | | | | |
| Episodes | 22642 | 25182 | 18772 | 26414 | 30251 | 20710 | 26018 | 29437 | 20722 |
| Individuals | 21818 | 24446 | 18208 | 25667 | 29480 | 20191 | 25847 | 29228 | 20612 |
| # Unique HCPs | 15545 | 18509 | 12741 | 17943 | 21735 | 14535 | 17950 | 21376 | 14460 |
| Total Cost | $19,574,941 | $20,622,560 | $19,582,054 | $22,168,601 | $23,868,328 | $21,222,948 | $21,334,742 | $22,658,013 | $20,491,857 |
| % Female | 51.4% | 51.7% | 52.3% | 51.0% | 50.6% | 52.2% | 51.6% | 51.4% | 51.9% |
| Age | 43 (32, 54) | 45 (33, 55) | 46 (33, 56) | 44 (32, 54) | 45 (33, 55) | 46 (33, 56) | 44 (32, 55) | 46 (33, 56) | 47 (34, 57) |
| ERG® Risk Score | 1.1 (0.5, 2.3) | 1.1 (0.5, 2.3) | 1.1 (0.6, 2.4) | 1.1 (0.5, 2.4) | 1.1 (0.5, 2.5) | 1.2 (0.6, 2.7) | 1.2 (0.6, 2.7) | 1.3 (0.6, 2.7) | 1.3 (0.6, 2.8) |
| % NHW | 63.2% (45.4%, 78.3%) | 69.6% (51.6%, 81.3%) | 64.7% (45.2%, 77.2%) | 63.8% (45.4%, 78.6%) | 70.0% (51.7%, 81.5%) | 65.0% (45.0%, 77.9%) | 63.1% (44.7%, 77.9%) | 70.4% (51.9%, 81.8%) | 64.6% (44.6%, 77.5%) |
| ADI | 43 (26, 62) | 46 (30, 62) | 16 (8, 33) | 45 (27, 64) | 47 (30, 63) | 18 (9, 38) | 44 (26, 63) | 46 (30, 62) | 17 (8, 36) |
| AGI | 74 (55, 108) | 70 (54, 100) | 89 (62, 151) | 71 (54, 104) | 70 (54, 96) | 85 (60, 142) | 73 (54, 106) | 70 (54, 99) | 87 (61, 143) |
| Episode Cost | $228 (90, 672) | $236 (92, 660) | $280 (97, 780) | $203 (68, 604) | $219 (74, 614) | $250 (68, 723) | $235 (80, 667) | $243 (86, 662) | $286 (79, 800) |
| # of HCP Seen | 1 (1, 2) | 1 (1, 2) | 1 (1, 2) | 1 (1, 2) | 1 (1, 2) | 1 (1, 2) | 1 (1, 2) | 1 (1, 2) | 1 (1, 2) |
| Episode Duration | 11 (1, 41) | 12 (1, 42) | 14 (1, 44) | 9 (1, 41) | 11 (1, 43) | 12 (1, 44) | 11 (1, 43) | 13 (1, 44) | 14 (1, 45) |
| | Medicare Advantage (MA) | | | | | | | | |
| Episodes | 52167 | 56931 | 31277 | 56383 | 61771 | 33312 | 63274 | 71268 | 36711 |
| Individuals | 50633 | 55122 | 30331 | 54990 | 60164 | 32468 | 63043 | 70989 | 36562 |
| # Unique HCPs | 27720 | 30421 | 19599 | 29900 | 33510 | 20726 | 32836 | 37161 | 22535 |
| Total Cost | $21,711,015 | $28,154,001 | $13,081,009 | $22,499,132 | $28,969,993 | $13,596,880 | $27,624,391 | $36,332,634 | $17,619,987 |
| % Female | 61.2% | 60.6% | 60.1% | 61.0% | 59.7% | 59.4% | 61.1% | 60.2% | 59.5% |
| Age | 71 (66, 77) | 72 (68, 78) | 72 (68, 79) | 71 (67, 77) | 73 (68, 78) | 73 (68, 79) | 72 (67, 78) | 73 (68, 79) | 73 (68, 79) |
| ERG® Risk Score | 0.9 (0.4, 1.7) | 1.0 (0.5, 1.7) | 0.9 (0.5, 1.6) | 1.0 (0.4, 1.8) | 1.0 (0.5, 1.8) | 1.0 (0.5, 1.8) | 1.1 (0.5, 1.9) | 1.1 (0.6, 1.9) | 1.1 (0.6, 1.8) |
| % NHW | 62.3% (41.3%, 78.4%) | 74.3% (55.7%, 84.7%) | 64.1% (42.2%, 81.1%) | 63.1% (42.1%, 78.6%) | 74.4% (56.2%, 84.9%) | 65.3% (43.0%, 81.6%) | 63.5% (42.8%, 78.9%) | 74.3% (56.5%, 84.7%) | 66.6% (44.2%, 82.1%) |
| ADI | 60 (42, 76) | 53 (38, 68) | 30 (16, 55) | 61 (43, 77) | 54 (38, 69) | 33 (17, 57) | 60 (42, 76) | 54 (38, 69) | 34 (17, 58) |
| AGI | 57 (46, 75) | 63 (51, 85) | 70 (54, 95) | 56 (46, 74) | 62 (51, 84) | 69 (54, 91) | 57 (46, 75) | 63 (51, 85) | 69 (54, 93) |
| Episode Cost | $115 (29, 358) | $159 (48, 454) | $111 (28, 358) | $97 (27, 323) | $139 (39, 411) | $91 (26, 330) | $125 (31, 397) | $180 (47, 495) | $127 (29, 425) |
| # of HCP Seen | 1 (1, 2) | 1 (1, 2) | 1 (1, 2) | 1 (1, 2) | 1 (1, 2) | 1 (1, 2) | 1 (1, 2) | 1 (1, 2) | 1 (1, 2) |
| Episode Duration | 14 (1, 48) | 16 (1, 49) | 16 (1, 49) | 11 (1, 49) | 14 (1, 50) | 13 (1, 50) | 15 (1, 50) | 17 (1, 50) | 17 (1, 51) |

Pre=Pre-COVID period, Early=first 12 months post-COVID, Late=13-24 months post-COVID

B25=25 least stringent States on 4/1/2020 and 4/1/2021, T25=25 most stringent States on 4/1/2020 and 4/1/2021, Mixed=In Bottom 25 on 4/1/2020 and Top 25 on 4/1/2021, or in Top 25 on 4/1/2020 and Bottom 25 on 4/1/2021

Within Pre-, Early and Late COVID periods compared to Top 25 baseline, cells in red are not significantly different - Mann-Whitney U test (p=0.05)
