## Supplement - Table 2 - Full Cohort for "Impact of the COVID-19 pandemic on low back pain management in commercially insured and Medicare Advantage cohorts. A retrospective cohort study"

**Supplement - Table 2 - Type of healthcare provider initially contacted for low back pain - Full Cohort**

| | | % of Episodes | | | RR and 95% CI Compared to Pre | |
|---|---|---|---|---|---|---|
| | | Pre-COVID | Early COVID | Late COVID | Early | Late |
| Commercial Insurance (CI) | | | | | | |
| Episodes | | 150761 | 158123 | 124958 | | |
| Non-Rx | DC | 23.6% | 24.6% | 24.1% | 1.04 (1.03, 1.05) | 1.02 (1.01, 1.03) |
| | PT | 3.7% | 3.1% | 3.4% | 0.86 (0.83, 0.89) | 0.94 (0.90, 0.97) |
| | LAc | 0.3% | 0.2% | 0.2% | 0.66 (0.57, 0.77) | 0.72 (0.61, 0.84) |
| Primary Care | PCP | 23.6% | 24.7% | 21.7% | 1.05 (1.03, 1.06) | 0.92 (0.91, 0.93) |
| | Nurse | 6.7% | 6.4% | 5.9% | 0.96 (0.93, 0.98) | 0.89 (0.87, 0.92) |
| | PA | 5.0% | 4.6% | 4.7% | 0.92 (0.89, 0.95) | 0.95 (0.92, 0.98) |
| | DO | 0.2% | 0.2% | 0.2% | 0.98 (0.84, 1.15) | 0.98 (0.83, 1.16) |
| Physician Specialist | OS | 7.2% | 7.4% | 8.2% | 1.03 (1.01, 1.06) | 1.14 (1.11, 1.17) |
| | PM | 4.4% | 3.2% | 2.4% | 0.72 (0.69, 0.75) | 0.54 (0.52, 0.56) |
| | PMR | 3.8% | 3.2% | 3.0% | 0.84 (0.81, 0.87) | 0.80 (0.77, 0.83) |
| | Neuro | 0.8% | 0.7% | 0.6% | 0.88 (0.81, 0.95) | 0.70 (0.64, 0.76) |
| | Rheum | 0.8% | 0.8% | 0.6% | 0.94 (0.87, 1.02) | 0.76 (0.69, 0.83) |
| | NS | 0.4% | 0.4% | 0.4% | 1.01 (0.91, 1.12) | 0.94 (0.83, 1.05) |
| | Oth | 1.9% | 2.2% | 1.9% | 1.13 (1.07, 1.19) | 1.00 (0.95, 1.05) |
| Emergency Medicine/ Urgent Care | Hosp | 8.4% | 8.3% | 10.2% | 1.00 (0.97, 1.02) | 1.22 (1.19, 1.25) |
| | Rad | 6.8% | 7.6% | 9.5% | 1.11 (1.08, 1.14) | 1.40 (1.36, 1.43) |
| | EM | 1.8% | 1.9% | 2.2% | 1.03 (0.98, 1.09) | 1.17 (1.11, 1.23) |
| | UC | 0.6% | 0.6% | 0.7% | 1.02 (0.93, 1.12) | 1.14 (1.04, 1.26) |
| Medicare Advantage (MA) | | | | | | |
| Episodes | | 530583 | 493573 | 368842 | | |
| Non-Rx | DC | 7.7% | 7.4% | 8.4% | 0.97 (0.95, 0.98) | 1.09 (1.08, 1.11) |
| | PT | 2.7% | 2.3% | 3.0% | 0.87 (0.85, 0.89) | 1.12 (1.09, 1.15) |
| | LAc | 0.1% | 0.1% | 0.1% | 0.96 (0.83, 1.12) | 1.69 (1.47, 1.95) |
| Primary Care | PCP | 36.3% | 39.7% | 34.2% | 1.09 (1.09, 1.10) | 0.94 (0.94, 0.95) |
| | Nurse | 7.8% | 7.8% | 6.8% | 1.00 (0.99, 1.01) | 0.87 (0.85, 0.88) |
| | PA | 4.9% | 4.7% | 4.5% | 0.94 (0.92, 0.96) | 0.92 (0.90, 0.93) |
| | DO | 0.1% | 0.1% | 0.1% | 0.99 (0.89, 1.11) | 1.04 (0.92, 1.17) |
| Physician Specialist | OS | 4.8% | 5.0% | 5.9% | 1.05 (1.03, 1.07) | 1.24 (1.22, 1.27) |
| | PM | 8.6% | 5.7% | 3.8% | 0.66 (0.65, 0.67) | 0.44 (0.43, 0.45) |
| | PMR | 3.5% | 2.7% | 2.4% | 0.76 (0.74, 0.77) | 0.67 (0.65, 0.69) |
| | Neuro | 0.9% | 0.9% | 0.6% | 0.96 (0.92, 1.00) | 0.71 (0.68, 0.75) |
| | Rheum | 0.9% | 0.9% | 0.8% | 0.98 (0.94, 1.02) | 0.81 (0.77, 0.84) |
| | NS | 0.4% | 0.4% | 0.3% | 0.89 (0.84, 0.95) | 0.80 (0.75, 0.86) |
| | Oth | 2.1% | 2.3% | 2.1% | 1.13 (1.11, 1.16) | 1.03 (1.00, 1.06) |
| Emergency Medicine/ Urgent Care | Hosp | 10.6% | 10.9% | 14.6% | 1.03 (1.02, 1.04) | 1.38 (1.37, 1.40) |
| | Rad | 7.3% | 7.9% | 10.6% | 1.08 (1.07, 1.10) | 1.46 (1.44, 1.48) |
| | EM | 1.1% | 1.1% | 1.4% | 0.96 (0.92, 0.99) | 1.27 (1.22, 1.32) |
| | UC | 0.3% | 0.3% | 0.4% | 1.01 (0.94, 1.09) | 1.31 (1.21, 1.41) |

Pre=Pre-COVID period, Early=first 12 months post-COVID, Late=13-24 months post-COVID

PCP=primary care physician, PA=physician assistant, DO=doctor of osteopathy, DC=doctor of chiropractic, PT=physical therapist, LAc=licensed acupuncturist, OS=orthopedic surgeon, PMR=physical medicine and rehabilitation, PM=pain management, Neuro=neurologist, Rheum=rheumatologist, NS=neurosurgeon, MD-Oth=other physician specialtists, Hosp=hospital, Rad=radiologist, EM=emergency medicine, UC=urgent care

For %, cells with red text indicate the type of HCP initially contacted was not signficantly different from pre-COVID period (Fisher's Exact p > .05)
For %, cells with black text indicate the type of HCP initially contacted was signficantly different from pre-COVID period (Fisher's Exact p > .05)
For risk ratio, cells in red indicate the confidence interval includes 1 and likelihood of HCP being initially contacted is not different than the pre-COVID reference
