## Supplement - Table 2 - Stringency for "Impact of the COVID-19 pandemic on low back pain management in commercially insured and Medicare Advantage cohorts. A retrospective cohort study"

**Supplement - Table 2 - Type of Health Care Provider Initially Contacted in the Pre-, Early and Late COVID Periods by Stringency of State Public Policy Response**

| | Type of HCP Initially Contacted | | | | | | | | | Risk Ratio and 95% Confidence Interval Compared to Top 25 Baseline | | | | | | Risk Ratio and 95% Confidence Interval Compared to Pre-COVID Baseline | | | | | |
|---|---|---|---|---|---|---|---|---|---|---|---|---|---|---|---|---|---|---|---|---|---|
| | Pre-COVID | | | Early COVID | | | Late COVID | | | Pre-COVID | | Early COVID | | Late COVID | | Early COVID | | | Late COVID | | |
| | B25 | Mixed | T25 | B25 | Mixed | T25 | B25 | Mixed | T25 | Bottom 25 | Mixed | Bottom 25 | Mixed | Bottom 25 | Mixed | Bottom 25 | Mixed | Top 25 | Bottom 25 | Mixed | Top 25 |
| | Commercial Insurance (CI) | | | | | | | | | | | | | | | | | | | | |
| PCP | 20.0% | 20.1% | 21.1% | 21.5% | 21.8% | 22.9% | 19.2% | 19.7% | 20.3% | 0.95 (0.91, 0.99) | 0.95 (0.92, 0.99) | 0.94 (0.91, 0.97) | 0.95 (0.92, 0.98) | 0.95 (0.91, 0.98) | 0.97 (0.94, 1.01) | 1.07 (1.04, 1.11) | 1.08 (1.05, 1.12) | 1.09 (1.05, 1.13) | 0.96 (0.93, 1.00) | 0.98 (0.95, 1.01) | 0.96 (0.92, 1.00) |
| Nurse | 6.1% | 6.1% | 4.2% | 6.3% | 6.3% | 4.3% | 6.2% | 6.3% | 4.1% | 1.43 (1.32, 1.56) | 1.44 (1.33, 1.57) | 1.48 (1.36, 1.60) | 1.48 (1.37, 1.60) | 1.51 (1.39, 1.63) | 1.51 (1.40, 1.64) | 1.04 (0.97, 1.12) | 1.04 (0.97, 1.11) | 1.01 (0.92, 1.11) | 1.03 (0.96, 1.10) | 1.02 (0.96, 1.09) | 0.98 (0.89, 1.07) |
| PA | 4.0% | 5.6% | 5.7% | 3.9% | 5.1% | 5.3% | 3.9% | 5.3% | 5.2% | 0.71 (0.65, 0.77) | 0.98 (0.90, 1.06) | 0.73 (0.67, 0.80) | 0.98 (0.91, 1.05) | 0.75 (0.69, 0.82) | 1.02 (0.95, 1.10) | 0.96 (0.88, 1.05) | 0.93 (0.86, 0.99) | 0.93 (0.85, 1.01) | 0.98 (0.90, 1.07) | 0.96 (0.90, 1.03) | 0.92 (0.85, 1.00) |
| DO | 0.2% | 0.2% | 0.2% | 0.2% | 0.2% | 0.3% | 0.1% | 0.2% | 0.2% | 0.81 (0.53, 1.24) | 1.01 (0.68, 1.49) | 0.76 (0.52, 1.10) | 0.73 (0.51, 1.05) | 0.58 (0.38, 0.90) | 1.00 (0.69, 1.44) | 1.12 (0.75, 1.68) | 0.88 (0.61, 1.25) | 1.20 (0.81, 1.78) | 0.73 (0.46, 1.14) | 1.00 (0.71, 1.42) | 1.01 (0.67, 1.53) |
| DC | 30.5% | 27.6% | 20.7% | 30.6% | 27.7% | 20.4% | 29.2% | 26.3% | 19.2% | 1.48 (1.43, 1.53) | 1.33 (1.29, 1.38) | 1.50 (1.45, 1.55) | 1.36 (1.32, 1.41) | 1.52 (1.47, 1.57) | 1.37 (1.32, 1.42) | 1.00 (0.97, 1.03) | 1.01 (0.98, 1.03) | 0.99 (0.95, 1.03) | 0.96 (0.93, 0.98) | 0.95 (0.93, 0.98) | 0.93 (0.89, 0.97) |
| PT | 3.0% | 2.9% | 6.2% | 2.5% | 2.4% | 5.0% | 2.7% | 2.9% | 4.7% | 0.48 (0.44, 0.53) | 0.46 (0.42, 0.51) | 0.49 (0.45, 0.55) | 0.49 (0.45, 0.54) | 0.59 (0.54, 0.65) | 0.62 (0.57, 0.68) | 0.83 (0.74, 0.92) | 0.85 (0.77, 0.94) | 0.80 (0.74, 0.87) | 0.92 (0.83, 1.02) | 1.01 (0.92, 1.11) | 0.75 (0.69, 0.81) |
| LAc | 0.1% | 0.2% | 0.5% | 0.0% | 0.1% | 0.3% | 0.0% | 0.1% | 0.5% | 0.11 (0.06, 0.20) | 0.41 (0.29, 0.57) | 0.15 (0.08, 0.27) | 0.43 (0.29, 0.62) | 0.10 (0.06, 0.18) | 0.28 (0.20, 0.40) | 0.86 (0.40, 1.85) | 0.68 (0.45, 1.01) | 0.64 (0.47, 0.88) | 0.87 (0.40, 1.88) | 0.66 (0.44, 0.99) | 0.96 (0.73, 1.27) |
| OS | 8.1% | 8.2% | 9.5% | 7.9% | 8.3% | 9.6% | 8.4% | 8.7% | 9.6% | 0.85 (0.80, 0.91) | 0.87 (0.82, 0.93) | 0.83 (0.78, 0.88) | 0.87 (0.82, 0.92) | 0.87 (0.83, 0.93) | 0.90 (0.86, 0.96) | 0.98 (0.92, 1.04) | 1.01 (0.95, 1.07) | 1.02 (0.96, 1.08) | 1.04 (0.98, 1.11) | 1.06 (1.00, 1.12) | 1.02 (0.96, 1.08) |
| PMR | 2.7% | 2.5% | 4.8% | 2.4% | 2.2% | 4.1% | 2.5% | 2.2% | 4.0% | 0.58 (0.52, 0.64) | 0.52 (0.47, 0.57) | 0.58 (0.52, 0.64) | 0.54 (0.49, 0.60) | 0.62 (0.56, 0.68) | 0.54 (0.49, 0.60) | 0.86 (0.77, 0.96) | 0.90 (0.81, 1.01) | 0.86 (0.79, 0.95) | 0.91 (0.82, 1.02) | 0.89 (0.80, 0.99) | 0.85 (0.78, 0.93) |
| PM | 1.7% | 1.5% | 2.0% | 1.8% | 1.6% | 2.0% | 1.7% | 1.6% | 1.8% | 0.86 (0.75, 0.99) | 0.76 (0.66, 0.87) | 0.91 (0.80, 1.04) | 0.78 (0.68, 0.89) | 0.95 (0.83, 1.09) | 0.92 (0.81, 1.06) | 1.04 (0.91, 1.19) | 1.01 (0.88, 1.16) | 0.98 (0.86, 1.13) | 0.96 (0.84, 1.10) | 1.07 (0.94, 1.22) | 0.88 (0.76, 1.01) |
| Neuro | 0.4% | 0.3% | 0.9% | 0.3% | 0.4% | 0.8% | 0.3% | 0.4% | 0.7% | 0.43 (0.33, 0.56) | 0.40 (0.31, 0.52) | 0.40 (0.31, 0.51) | 0.53 (0.42, 0.66) | 0.40 (0.30, 0.53) | 0.50 (0.39, 0.65) | 0.87 (0.64, 1.17) | 1.24 (0.95, 1.62) | 0.94 (0.76, 1.16) | 0.75 (0.55, 1.02) | 1.01 (0.76, 1.34) | 0.80 (0.64, 1.00) |
| Rheum | 0.4% | 0.4% | 0.6% | 0.5% | 0.5% | 0.6% | 0.4% | 0.5% | 0.6% | 0.67 (0.51, 0.88) | 0.66 (0.51, 0.86) | 0.80 (0.62, 1.04) | 0.86 (0.67, 1.09) | 0.76 (0.59, 0.99) | 0.89 (0.69, 1.13) | 1.11 (0.85, 1.45) | 1.20 (0.93, 1.54) | 0.92 (0.71, 1.19) | 1.05 (0.80, 1.38) | 1.24 (0.96, 1.59) | 0.92 (0.71, 1.19) |
| NS | 0.3% | 0.4% | 0.3% | 0.4% | 0.3% | 0.2% | 0.3% | 0.4% | 0.3% | 0.97 (0.67, 1.40) | 1.46 (1.05, 2.04) | 1.65 (1.17, 2.33) | 1.50 (1.06, 2.11) | 1.19 (0.85, 1.66) | 1.54 (1.13, 2.11) | 1.40 (1.02, 1.91) | 0.84 (0.64, 1.10) | 0.82 (0.56, 1.21) | 1.19 (0.86, 1.65) | 1.03 (0.79, 1.33) | 0.97 (0.67, 1.42) |
| MD Oth | 1.3% | 1.4% | 1.6% | 1.6% | 1.6% | 2.2% | 1.4% | 1.5% | 1.9% | 0.80 (0.68, 0.93) | 0.85 (0.73, 0.99) | 0.72 (0.64, 0.82) | 0.71 (0.63, 0.81) | 0.76 (0.66, 0.87) | 0.80 (0.70, 0.92) | 1.29 (1.11, 1.49) | 1.18 (1.03, 1.36) | 1.41 (1.22, 1.63) | 1.13 (0.97, 1.31) | 1.11 (0.96, 1.28) | 1.18 (1.02, 1.37) |
| Hosp | 9.4% | 11.0% | 10.2% | 8.6% | 9.8% | 10.3% | 10.3% | 11.5% | 11.2% | 0.92 (0.86, 0.97) | 1.07 (1.02, 1.13) | 0.83 (0.79, 0.88) | 0.95 (0.90, 1.00) | 0.92 (0.88, 0.97) | 1.03 (0.98, 1.09) | 0.92 (0.87, 0.97) | 0.89 (0.85, 0.93) | 1.01 (0.95, 1.07) | 1.10 (1.04, 1.16) | 1.05 (1.00, 1.10) | 1.09 (1.03, 1.15) |
| Rad | 8.2% | 8.1% | 8.2% | 7.9% | 8.2% | 8.6% | 9.3% | 8.9% | 12.5% | 1.00 (0.94, 1.07) | 0.99 (0.93, 1.05) | 0.92 (0.87, 0.98) | 0.95 (0.90, 1.01) | 0.75 (0.71, 0.79) | 0.71 (0.68, 0.75) | 0.97 (0.91, 1.03) | 1.02 (0.96, 1.07) | 1.05 (0.99, 1.12) | 1.14 (1.07, 1.21) | 1.10 (1.04, 1.16) | 1.52 (1.44, 1.62) |
| EM | 2.6% | 2.6% | 2.4% | 2.7% | 2.5% | 2.2% | 2.9% | 2.6% | 2.3% | 1.06 (0.94, 1.20) | 1.07 (0.95, 1.20) | 1.24 (1.11, 1.39) | 1.12 (1.00, 1.26) | 1.22 (1.09, 1.37) | 1.10 (0.98, 1.23) | 1.05 (0.94, 1.17) | 0.94 (0.85, 1.05) | 0.90 (0.79, 1.02) | 1.11 (0.99, 1.23) | 0.99 (0.90, 1.10) | 0.96 (0.85, 1.09) |
| UC | 1.1% | 0.8% | 0.9% | 0.9% | 0.8% | 0.9% | 0.9% | 0.8% | 0.9% | 1.21 (1.00, 1.48) | 0.97 (0.79, 1.19) | 0.98 (0.81, 1.20) | 0.88 (0.72, 1.07) | 1.01 (0.83, 1.23) | 0.89 (0.73, 1.08) | 0.81 (0.67, 0.97) | 0.90 (0.75, 1.09) | 0.99 (0.81, 1.23) | 0.83 (0.70, 1.00) | 0.92 (0.76, 1.10) | 1.00 (0.81, 1.23) |
| | Medicare Advantage (MA) | | | | | | | | | | | | | | | | | | | | |
| PCP | 34.1% | 27.4% | 34.7% | 37.8% | 30.6% | 38.3% | 32.2% | 25.6% | 32.5% | 0.98 (0.96, 1.00) | 0.79 (0.77, 0.81) | 0.99 (0.97, 1.00) | 0.80 (0.78, 0.81) | 0.99 (0.97, 1.01) | 0.79 (0.77, 0.80) | 1.11 (1.09, 1.13) | 1.11 (1.10, 1.14) | 1.10 (1.08, 1.13) | 0.94 (0.93, 0.96) | 0.93 (0.92, 0.95) | 0.94 (0.92, 0.96) |
| Nurse | 6.9% | 5.9% | 5.2% | 7.4% | 6.5% | 5.6% | 7.2% | 6.0% | 4.9% | 1.34 (1.26, 1.42) | 1.15 (1.09, 1.22) | 1.33 (1.26, 1.40) | 1.17 (1.11, 1.23) | 1.47 (1.40, 1.55) | 1.24 (1.18, 1.31) | 1.07 (1.03, 1.12) | 1.10 (1.05, 1.15) | 1.08 (1.01, 1.15) | 1.04 (1.00, 1.08) | 1.02 (0.97, 1.06) | 0.94 (0.88, 1.01) |
| PA | 4.2% | 4.4% | 5.1% | 3.9% | 4.6% | 5.0% | 4.1% | 4.9% | 5.1% | 0.81 (0.76, 0.86) | 0.85 (0.80, 0.91) | 0.78 (0.73, 0.83) | 0.91 (0.86, 0.97) | 0.81 (0.76, 0.85) | 0.96 (0.91, 1.01) | 0.94 (0.89, 1.00) | 1.05 (0.99, 1.10) | 0.98 (0.92, 1.05) | 0.99 (0.94, 1.05) | 1.11 (1.06, 1.17) | 1.00 (0.93, 1.06) |
| DO | 0.1% | 0.1% | 0.1% | 0.1% | 0.1% | 0.1% | 0.1% | 0.1% | 0.1% | 1.03 (0.70, 1.53) | 1.09 (0.74, 1.59) | 0.91 (0.60, 1.38) | 1.15 (0.78, 1.70) | 0.91 (0.65, 1.29) | 0.95 (0.68, 1.33) | 0.76 (0.54, 1.09) | 0.92 (0.67, 1.26) | 0.87 (0.56, 1.36) | 0.98 (0.71, 1.35) | 0.97 (0.72, 1.31) | 1.11 (0.73, 1.67) |
| DC | 8.0% | 11.6% | 8.7% | 7.3% | 10.4% | 7.7% | 8.2% | 10.3% | 8.6% | 0.92 (0.88, 0.96) | 1.33 (1.27, 1.39) | 0.95 (0.90, 0.99) | 1.35 (1.29, 1.41) | 0.96 (0.92, 1.01) | 1.20 (1.15, 1.25) | 0.91 (0.87, 0.95) | 0.90 (0.87, 0.93) | 0.88 (0.84, 0.93) | 1.03 (0.99, 1.07) | 0.89 (0.86, 0.92) | 0.98 (0.93, 1.03) |
| PT | 2.8% | 3.9% | 3.9% | 2.2% | 3.0% | 3.0% | 2.7% | 3.7% | 3.7% | 0.73 (0.68, 0.78) | 1.00 (0.93, 1.07) | 0.73 (0.68, 0.80) | 1.01 (0.94, 1.09) | 0.71 (0.66, 0.76) | 0.98 (0.92, 1.04) | 0.78 (0.72, 0.84) | 0.78 (0.73, 0.83) | 0.77 (0.71, 0.83) | 0.93 (0.87, 1.00) | 0.93 (0.88, 0.99) | 0.95 (0.88, 1.03) |
| LAc | 0.0% | 0.0% | 0.3% | 0.0% | 0.0% | 0.2% | 0.0% | 0.1% | 0.3% | 0.07 (0.04, 0.14) | 0.14 (0.09, 0.22) | 0.12 (0.07, 0.22) | 0.21 (0.13, 0.34) | 0.09 (0.05, 0.14) | 0.21 (0.15, 0.29) | 1.20 (0.53, 2.74) | 1.10 (0.61, 1.96) | 0.71 (0.51, 0.99) | 1.48 (0.69, 3.21) | 1.83 (1.09, 3.05) | 1.23 (0.93, 1.63) |
| OS | 6.8% | 7.1% | 4.7% | 6.8% | 6.9% | 4.6% | 7.3% | 7.3% | 4.9% | 1.45 (1.37, 1.54) | 1.51 (1.42, 1.60) | 1.46 (1.38, 1.55) | 1.49 (1.40, 1.57) | 1.48 (1.40, 1.56) | 1.48 (1.40, 1.56) | 1.00 (0.96, 1.04) | 0.98 (0.94, 1.02) | 0.99 (0.92, 1.06) | 1.07 (1.02, 1.11) | 1.03 (0.99, 1.07) | 1.05 (0.98, 1.12) |
| PMR | 2.4% | 2.1% | 2.5% | 2.0% | 2.0% | 2.1% | 2.1% | 2.0% | 2.1% | 0.96 (0.87, 1.04) | 0.84 (0.77, 0.91) | 0.95 (0.86, 1.04) | 0.95 (0.87, 1.05) | 0.99 (0.91, 1.09) | 0.98 (0.90, 1.07) | 0.82 (0.76, 0.89) | 0.94 (0.87, 1.02) | 0.83 (0.75, 0.91) | 0.86 (0.80, 0.93) | 0.97 (0.90, 1.04) | 0.83 (0.75, 0.91) |
| PM | 3.3% | 3.0% | 2.1% | 3.1% | 2.6% | 2.0% | 3.1% | 2.6% | 1.8% | 1.54 (1.41, 1.69) | 1.42 (1.30, 1.55) | 1.56 (1.43, 1.71) | 1.32 (1.21, 1.44) | 1.76 (1.61, 1.92) | 1.45 (1.33, 1.59) | 0.95 (0.89, 1.02) | 0.87 (0.82, 0.93) | 0.94 (0.85, 1.05) | 0.94 (0.89, 1.01) | 0.85 (0.80, 0.91) | 0.83 (0.75, 0.92) |
| Neuro | 0.5% | 0.5% | 0.6% | 0.6% | 0.6% | 0.6% | 0.5% | 0.5% | 0.5% | 0.75 (0.63, 0.91) | 0.79 (0.66, 0.95) | 0.93 (0.78, 1.11) | 1.01 (0.85, 1.20) | 0.89 (0.75, 1.07) | 0.92 (0.77, 1.10) | 1.14 (0.96, 1.34) | 1.18 (1.01, 1.38) | 0.92 (0.76, 1.12) | 0.98 (0.83, 1.16) | 0.97 (0.83, 1.13) | 0.83 (0.68, 1.01) |
| Rheum | 0.7% | 0.9% | 0.8% | 0.7% | 0.8% | 0.6% | 0.6% | 0.7% | 0.6% | 0.91 (0.78, 1.07) | 1.14 (0.98, 1.33) | 1.13 (0.96, 1.34) | 1.32 (1.13, 1.55) | 1.05 (0.88, 1.24) | 1.28 (1.09, 1.51) | 1.01 (0.87, 1.16) | 0.94 (0.83, 1.06) | 0.81 (0.67, 0.98) | 0.84 (0.72, 0.96) | 0.82 (0.73, 0.93) | 0.73 (0.61, 0.88) |
| NS | 0.4% | 0.3% | 0.1% | 0.3% | 0.3% | 0.2% | 0.3% | 0.3% | 0.2% | 2.97 (2.10, 4.18) | 2.63 (1.87, 3.72) | 2.03 (1.49, 2.75) | 1.77 (1.30, 2.41) | 1.98 (1.48, 2.65) | 1.71 (1.28, 2.30) | 0.87 (0.71, 1.07) | 0.86 (0.70, 1.05) | 1.28 (0.84, 1.93) | 0.85 (0.69, 1.03) | 0.82 (0.67, 1.01) | 1.27 (0.84, 1.90) |
| MD Oth | 2.0% | 1.6% | 1.9% | 2.3% | 2.1% | 2.4% | 2.1% | 1.7% | 2.1% | 1.07 (0.97, 1.18) | 0.88 (0.79, 0.98) | 0.97 (0.89, 1.05) | 0.86 (0.79, 0.94) | 0.99 (0.90, 1.08) | 0.82 (0.75, 0.89) | 1.17 (1.08, 1.27) | 1.27 (1.17, 1.38) | 1.30 (1.17, 1.45) | 1.04 (0.96, 1.13) | 1.06 (0.97, 1.15) | 1.14 (1.02, 1.26) |
| Hosp | 15.1% | 16.4% | 14.4% | 13.6% | 15.1% | 13.8% | 16.1% | 18.0% | 16.7% | 1.05 (1.01, 1.09) | 1.14 (1.10, 1.18) | 0.98 (0.95, 1.02) | 1.09 (1.05, 1.12) | 0.96 (0.94, 0.99) | 1.08 (1.05, 1.11) | 0.90 (0.88, 0.93) | 0.92 (0.90, 0.94) | 0.96 (0.93, 1.00) | 1.07 (1.04, 1.10) | 1.10 (1.07, 1.13) | 1.16 (1.12, 1.20) |
| Rad | 10.2% | 12.6% | 12.1% | 9.6% | 12.5% | 11.6% | 11.1% | 14.3% | 13.4% | 0.84 (0.81, 0.88) | 1.04 (1.00, 1.08) | 0.82 (0.79, 0.86) | 1.07 (1.03, 1.11) | 0.82 (0.80, 0.85) | 1.06 (1.03, 1.10) | 0.94 (0.90, 0.97) | 0.99 (0.96, 1.02) | 0.96 (0.92, 1.00) | 1.08 (1.05, 1.12) | 1.13 (1.10, 1.17) | 1.11 (1.07, 1.15) |
| EM | 2.2% | 1.7% | 1.9% | 1.9% | 1.6% | 1.4% | 2.0% | 1.7% | 1.8% | 1.14 (1.03, 1.26) | 0.89 (0.81, 0.99) | 1.35 (1.21, 1.50) | 1.12 (1.00, 1.24) | 1.14 (1.04, 1.25) | 0.95 (0.86, 1.04) | 0.88 (0.81, 0.96) | 0.93 (0.85, 1.01) | 0.74 (0.66, 0.84) | 0.94 (0.87, 1.02) | 1.00 (0.92, 1.08) | 0.94 (0.84, 1.05) |
| UC | 0.4% | 0.4% | 0.8% | 0.4% | 0.3% | 0.7% | 0.4% | 0.3% | 0.7% | 0.42 (0.35, 0.51) | 0.46 (0.38, 0.54) | 0.53 (0.44, 0.64) | 0.45 (0.37, 0.55) | 0.53 (0.45, 0.64) | 0.48 (0.40, 0.57) | 1.03 (0.84, 1.25) | 0.80 (0.66, 0.98) | 0.81 (0.68, 0.96) | 1.07 (0.88, 1.30) | 0.88 (0.73, 1.06) | 0.84 (0.71, 1.00) |

Pre=Pre-COVID period, Early=first 12 months post-COVID, Late=13-24 months post-COVID

B25=Bottom 25 least stringent States on 4/1/2020 and 4/1/2021, T25=Top 25 most stringent States on 4/1/2020 and 4/1/2021, Mixed=In Bottom 25 on 4/1/2020 and Top 25 on 4/1/2021, or in Top 25 on 4/1/2020 and Bottom 25 on 4/1/2021

PCP=primary care physician, PA=physician assistant, DO=doctor of osteopathy, DC=doctor of chiropractic, PT=physical therapist, LAc=licensed acupuncturist, OS=orthopedic surgeon, PMR=physical medicine and rehabilitation, PM=pain management, Neuro=neurologist, Rheum=rheumatologist, NS=neurosurgeon, MD-Oth=other physician specialtists, Hosp=hospital, Rad=radiologist, EM=emergency medicine, UC=urgent care

For %, cells with red text indicate the type of HCP initially contacted was not signficantly different from pre-COVID period (Fisher's Exact p > .05)

For %, cells with black text indicate the type of HCP initially contacted was signficantly different from pre-COVID period (Fisher's Exact p > .05)

For risk ratio, cells in red indicate the confidence interval includes 1 and likelihood of HCP being initially contacted is not different than the pre-COVID reference
