## Supplement - Table 3 - Full Cohort for "Impact of the COVID-19 pandemic on low back pain management in commercially insured and Medicare Advantage cohorts. A retrospective cohort study"

| Supplement - Table 3 - Type of healthcare services provided for low back pain - Full Cohort | | | | | | |
|---|---|---|---|---|---|---|
| | | % of Episodes | | | RR and 95% CI Compared to Pre | |
| | | Pre-COVID | Early COVID | Late COVID | Early | Late |
| Commercial Insurance (CI) | | | | | | |
| Episodes - At Least 1 Service | | 150705 | 158069 | 124913 | | |
| First Line | Manipulation - Chiropractic | 27.6% | 28.2% | 26.9% | 1.02 (1.01, 1.03) | 0.98 (0.96, 0.99) |
| | Active Care | 26.5% | 25.5% | 25.9% | 0.96 (0.95, 0.97) | 0.98 (0.97, 0.99) |
| | Manual Therapy | 17.8% | 16.4% | 16.3% | 0.92 (0.91, 0.94) | 0.92 (0.90, 0.93) |
| | Passive Therapy | 15.8% | 14.8% | 13.5% | 0.94 (0.92, 0.95) | 0.86 (0.84, 0.87) |
| | Acupuncture | 0.8% | 0.6% | 0.6% | 0.70 (0.65, 0.77) | 0.71 (0.65, 0.77) |
| | Manipulation - Osteopathic | 0.8% | 0.7% | 0.7% | 0.94 (0.87, 1.02) | 0.84 (0.77, 0.92) |
| Second Line | Imaging - Radiography | 36.8% | 37.4% | 41.1% | 1.02 (1.01, 1.03) | 1.12 (1.11, 1.13) |
| | Rx - NSAID | 31.9% | 30.5% | 27.8% | 0.96 (0.95, 0.97) | 0.87 (0.86, 0.88) |
| | Rx - Muscle Relaxant | 24.7% | 23.7% | 21.5% | 0.96 (0.95, 0.97) | 0.87 (0.86, 0.88) |
| | Imaging - MRI | 17.1% | 17.5% | 18.1% | 1.02 (1.01, 1.04) | 1.06 (1.04, 1.08) |
| | Rx Oral Steroid | 13.7% | 12.1% | 10.5% | 0.88 (0.86, 0.89) | 0.77 (0.75, 0.78) |
| | Rx Gabapentin | 4.2% | 3.6% | 2.9% | 0.86 (0.83, 0.89) | 0.69 (0.66, 0.72) |
| | Rx - Other | 15.8% | 14.1% | 11.2% | 0.89 (0.88, 0.91) | 0.71 (0.69, 0.72) |
| Third Line | Rx Opioid | 21.0% | 17.1% | 13.3% | 0.81 (0.80, 0.82) | 0.63 (0.62, 0.65) |
| | Spinal Injection | 13.7% | 12.4% | 10.8% | 0.90 (0.89, 0.92) | 0.78 (0.77, 0.80) |
| | Imaging - CT | 3.4% | 3.3% | 3.4% | 0.97 (0.94, 1.01) | 1.00 (0.96, 1.04) |
| | Surgery | 2.7% | 2.5% | 1.8% | 0.93 (0.89, 0.98) | 0.67 (0.64, 0.71) |
| Medicare Advantage (MA) | | | | | | |
| Episodes - At Least 1 Service | | 530269 | 493163 | 368380 | | |
| First Line | Manipulation - Chiropractic | 11.1% | 11.0% | 11.5% | 1.00 (0.98, 1.01) | 1.04 (1.03, 1.05) |
| | Active Care | 16.2% | 16.1% | 17.7% | 0.99 (0.98, 1.00) | 1.10 (1.09, 1.11) |
| | Manual Therapy | 8.2% | 7.9% | 8.6% | 0.98 (0.96, 0.99) | 1.05 (1.04, 1.07) |
| | Passive Therapy | 3.1% | 2.8% | 2.7% | 0.90 (0.88, 0.92) | 0.87 (0.84, 0.89) |
| | Acupuncture | 0.4% | 0.4% | 0.5% | 1.15 (1.08, 1.22) | 1.42 (1.33, 1.51) |
| | Manipulation - Osteopathic | 0.4% | 0.4% | 0.4% | 0.95 (0.89, 1.01) | 0.87 (0.81, 0.93) |
| Second Line | Imaging - Radiography | 33.7% | 34.4% | 38.5% | 1.02 (1.01, 1.02) | 1.14 (1.14, 1.15) |
| | Rx - NSAID | 32.4% | 30.4% | 25.9% | 0.94 (0.93, 0.94) | 0.80 (0.79, 0.81) |
| | Rx - Muscle Relaxant | 21.5% | 19.6% | 16.6% | 0.91 (0.90, 0.92) | 0.77 (0.77, 0.78) |
| | Imaging - MRI | 21.1% | 22.2% | 23.6% | 1.05 (1.04, 1.06) | 1.12 (1.11, 1.13) |
| | Rx Oral Steroid | 10.4% | 8.9% | 7.7% | 0.86 (0.85, 0.87) | 0.74 (0.73, 0.75) |
| | Rx Gabapentin | 5.0% | 4.4% | 3.0% | 0.87 (0.86, 0.89) | 0.60 (0.58, 0.61) |
| | Rx - Other | 26.9% | 26.8% | 20.3% | 1.00 (0.99, 1.00) | 0.76 (0.75, 0.76) |
| Third Line | Rx Opioid | 40.5% | 32.6% | 24.1% | 0.80 (0.80, 0.81) | 0.59 (0.59, 0.60) |
| | Spinal Injection | 21.6% | 19.7% | 17.1% | 0.91 (0.90, 0.92) | 0.79 (0.79, 0.80) |
| | Imaging - CT | 7.9% | 8.0% | 8.1% | 1.01 (1.00, 1.03) | 1.03 (1.02, 1.05) |
| | Surgery | 2.9% | 2.7% | 2.0% | 0.93 (0.91, 0.95) | 0.67 (0.66, 0.69) |

Pre=Pre-COVID period, Early=first 12 months post-COVID, Late=13-24 months post-COVID

For %, cells with red text indicate the type of HCP initially contacted was not signficantly different from pre-COVID period (Fisher's Exact p > .05)
For %, cells with black text indicate the type of HCP initially contacted was signficantly different from pre-COVID period (Fisher's Exact p > .05)
For risk ratio, cells in red indicate the confidence interval includes 1 and likelihood of HCP being initially contacted is not different than the pre-COVID reference
