## Supplement - Table 3 - Stringency for "Impact of the COVID-19 pandemic on low back pain management in commercially insured and Medicare Advantage cohorts. A retrospective cohort study"

**Supplement - Table 3 - Services Provided for LBP in the Pre-, Early and Late COVID Periods by Stringency of State Public Policy Response**

| | % of LBP Episodes Including Service | | | | | | | | | Risk Ratio and 95% Confidence Interval Compared to Top 25 Baseline | | | | | | Risk Ratio and 95% Confidence Interval Compared to Pre-COVID Baseline | | | | | |
|---|---|---|---|---|---|---|---|---|---|---|---|---|---|---|---|---|---|---|---|---|---|
| | Pre-COVID | | | Early COVID | | | Late COVID | | | Pre-COVID | | Early COVID | | Late COVID | | Early COVID | | | Late COVID | | |
| | B25 | Mixed | T25 | B25 | Mixed | T25 | B25 | Mixed | T25 | Bottom 25 | Mixed | Bottom 25 | Mixed | Bottom 25 | Mixed | Bottom 25 | Mixed | Top 25 | Bottom 25 | Mixed | Top 25 |
| Commercial | | | | | | | | | | | | | | | | | | | | | |
| At least one | 22642 | 25182 | 18772 | 26414 | 30251 | 20710 | 26018 | 29437 | 20722 | | | | | | | | | | | | |
| AC | 23.9% | 20.0% | 24.9% | 21.5% | 18.8% | 21.0% | 23.2% | 19.9% | 21.9% | 0.96 (0.93, 0.99) | 0.80 (0.78, 0.83) | 1.02 (0.98, 1.06) | 0.89 (0.86, 0.93) | 1.06 (1.03, 1.10) | 0.91 (0.88, 0.94) | 0.90 (0.87, 0.93) | 0.94 (0.91, 0.97) | 0.85 (0.82, 0.88) | 0.97 (0.94, 1.00) | 0.99 (0.96, 1.03) | 0.88 (0.85, 0.91) |
| MT | 13.2% | 12.8% | 18.4% | 11.4% | 11.5% | 14.7% | 11.7% | 11.8% | 15.3% | 0.72 (0.68, 0.75) | 0.70 (0.67, 0.73) | 0.77 (0.74, 0.81) | 0.78 (0.74, 0.81) | 0.77 (0.73, 0.80) | 0.77 (0.74, 0.81) | 0.86 (0.82, 0.91) | 0.90 (0.86, 0.94) | 0.80 (0.77, 0.84) | 0.89 (0.85, 0.93) | 0.92 (0.88, 0.96) | 0.83 (0.80, 0.87) |
| CMT | 29.2% | 28.5% | 21.8% | 29.2% | 28.7% | 21.4% | 28.3% | 27.6% | 20.2% | 1.34 (1.29, 1.38) | 1.31 (1.26, 1.35) | 1.37 (1.32, 1.41) | 1.34 (1.30, 1.39) | 1.40 (1.36, 1.45) | 1.36 (1.32, 1.41) | 1.00 (0.97, 1.03) | 1.01 (0.98, 1.04) | 0.98 (0.95, 1.02) | 0.97 (0.95, 1.00) | 0.97 (0.94, 1.00) | 0.93 (0.89, 0.96) |
| PT | 13.9% | 14.4% | 12.8% | 12.9% | 13.0% | 10.9% | 12.4% | 11.8% | 10.4% | 1.09 (1.04, 1.15) | 1.13 (1.07, 1.18) | 1.18 (1.13, 1.25) | 1.20 (1.14, 1.26) | 1.19 (1.13, 1.25) | 1.13 (1.07, 1.19) | 0.93 (0.89, 0.97) | 0.91 (0.87, 0.94) | 0.85 (0.81, 0.90) | 0.89 (0.85, 0.93) | 0.82 (0.79, 0.86) | 0.82 (0.77, 0.86) |
| Acu | 0.2% | 0.4% | 1.1% | 0.2% | 0.3% | 0.7% | 0.2% | 0.3% | 0.9% | 0.20 (0.15, 0.27) | 0.33 (0.26, 0.42) | 0.30 (0.22, 0.40) | 0.38 (0.29, 0.50) | 0.26 (0.19, 0.35) | 0.40 (0.31, 0.51) | 0.94 (0.64, 1.38) | 0.74 (0.55, 1.00) | 0.64 (0.52, 0.79) | 1.03 (0.70, 1.50) | 0.96 (0.72, 1.27) | 0.79 (0.65, 0.97) |
| OMT | 0.6% | 0.7% | 1.0% | 0.4% | 0.5% | 1.1% | 0.5% | 0.5% | 0.8% | 0.64 (0.52, 0.80) | 0.67 (0.54, 0.82) | 0.40 (0.32, 0.51) | 0.43 (0.35, 0.53) | 0.58 (0.46, 0.74) | 0.57 (0.45, 0.71) | 0.68 (0.53, 0.87) | 0.70 (0.56, 0.88) | 1.08 (0.89, 1.32) | 0.76 (0.60, 0.97) | 0.71 (0.57, 0.89) | 0.84 (0.68, 1.03) |
| Image - Rad | 37.4% | 39.1% | 35.1% | 35.6% | 36.8% | 35.3% | 39.1% | 40.2% | 39.6% | 1.07 (1.04, 1.09) | 1.11 (1.09, 1.14) | 1.01 (0.98, 1.03) | 1.04 (1.02, 1.07) | 0.99 (0.97, 1.01) | 1.02 (0.99, 1.04) | 0.95 (0.93, 0.97) | 0.94 (0.92, 0.96) | 1.01 (0.98, 1.03) | 1.05 (1.02, 1.07) | 1.03 (1.01, 1.05) | 1.13 (1.10, 1.16) |
| Image - MRI | 10.3% | 10.9% | 10.4% | 9.7% | 10.1% | 10.9% | 11.4% | 11.4% | 14.2% | 0.99 (0.94, 1.05) | 1.04 (0.99, 1.10) | 0.90 (0.85, 0.95) | 0.93 (0.88, 0.98) | 0.80 (0.77, 0.84) | 0.81 (0.77, 0.84) | 0.94 (0.90, 1.00) | 0.93 (0.88, 0.97) | 1.04 (0.99, 1.11) | 1.10 (1.05, 1.16) | 1.05 (1.00, 1.10) | 1.36 (1.29, 1.44) |
| Rx - NSAID | 21.5% | 21.8% | 24.4% | 22.4% | 22.4% | 24.0% | 22.2% | 22.1% | 22.2% | 0.88 (0.85, 0.91) | 0.89 (0.86, 0.92) | 0.93 (0.90, 0.96) | 0.93 (0.90, 0.96) | 1.00 (0.97, 1.04) | 1.00 (0.96, 1.03) | 1.04 (1.00, 1.07) | 1.03 (1.00, 1.06) | 0.98 (0.95, 1.02) | 1.03 (1.00, 1.07) | 1.02 (0.98, 1.05) | 0.91 (0.88, 0.94) |
| Rx - MM Rlx | 20.6% | 21.6% | 20.4% | 20.9% | 21.3% | 20.5% | 19.0% | 20.0% | 19.2% | 1.01 (0.97, 1.05) | 1.06 (1.02, 1.10) | 1.02 (0.99, 1.06) | 1.04 (1.00, 1.08) | 0.99 (0.95, 1.03) | 1.04 (1.00, 1.08) | 1.02 (0.98, 1.06) | 0.98 (0.95, 1.02) | 1.00 (0.96, 1.04) | 0.92 (0.89, 0.96) | 0.93 (0.90, 0.96) | 0.94 (0.90, 0.98) |
| Rx - Steroid | 10.7% | 12.2% | 10.4% | 9.9% | 11.1% | 8.8% | 9.1% | 9.5% | 7.8% | 1.03 (0.98, 1.09) | 1.18 (1.12, 1.24) | 1.13 (1.06, 1.19) | 1.26 (1.20, 1.33) | 1.16 (1.09, 1.23) | 1.21 (1.14, 1.29) | 0.92 (0.87, 0.97) | 0.91 (0.87, 0.95) | 0.85 (0.80, 0.90) | 0.85 (0.80, 0.89) | 0.77 (0.74, 0.81) | 0.75 (0.71, 0.80) |
| Rx - Gaba | 1.4% | 1.4% | 1.8% | 1.5% | 1.6% | 2.1% | 1.5% | 1.4% | 2.1% | 0.76 (0.65, 0.89) | 0.80 (0.69, 0.92) | 0.73 (0.64, 0.83) | 0.75 (0.66, 0.85) | 0.71 (0.62, 0.81) | 0.67 (0.59, 0.77) | 1.11 (0.96, 1.28) | 1.09 (0.95, 1.24) | 1.16 (1.01, 1.33) | 1.06 (0.92, 1.23) | 0.97 (0.84, 1.11) | 1.14 (0.99, 1.31) |
| Rx - Oth | 5.5% | 6.3% | 6.4% | 6.1% | 6.9% | 7.0% | 5.7% | 6.7% | 6.3% | 0.87 (0.80, 0.93) | 0.98 (0.91, 1.06) | 0.87 (0.81, 0.93) | 1.00 (0.93, 1.06) | 0.90 (0.84, 0.97) | 1.06 (0.99, 1.13) | 1.10 (1.02, 1.18) | 1.11 (1.04, 1.18) | 1.09 (1.01, 1.18) | 1.03 (0.96, 1.11) | 1.07 (1.00, 1.14) | 0.99 (0.92, 1.07) |
| Rx - Opioid | 10.1% | 9.4% | 8.4% | 9.8% | 9.8% | 8.8% | 9.5% | 8.7% | 7.8% | 1.19 (1.12, 1.27) | 1.12 (1.05, 1.19) | 1.11 (1.05, 1.18) | 1.12 (1.06, 1.18) | 1.21 (1.14, 1.28) | 1.11 (1.04, 1.18) | 0.97 (0.92, 1.02) | 1.04 (0.99, 1.09) | 1.04 (0.97, 1.11) | 0.94 (0.89, 0.99) | 0.92 (0.88, 0.97) | 0.93 (0.87, 0.99) |
| Spinal Inj | 5.0% | 4.8% | 6.3% | 5.0% | 4.7% | 6.2% | 5.1% | 5.3% | 6.5% | 0.79 (0.73, 0.86) | 0.77 (0.71, 0.83) | 0.81 (0.75, 0.87) | 0.76 (0.70, 0.82) | 0.78 (0.72, 0.84) | 0.81 (0.75, 0.87) | 1.00 (0.93, 1.08) | 0.97 (0.90, 1.04) | 0.99 (0.91, 1.06) | 1.02 (0.94, 1.10) | 1.08 (1.01, 1.17) | 1.04 (0.96, 1.12) |
| Image - CT | 2.7% | 2.8% | 1.7% | 2.7% | 2.7% | 2.0% | 3.4% | 2.9% | 2.2% | 1.53 (1.34, 1.75) | 1.63 (1.43, 1.85) | 1.37 (1.21, 1.54) | 1.36 (1.21, 1.53) | 1.54 (1.38, 1.72) | 1.34 (1.20, 1.50) | 1.02 (0.92, 1.14) | 0.96 (0.87, 1.06) | 1.14 (0.99, 1.32) | 1.26 (1.13, 1.39) | 1.03 (0.93, 1.14) | 1.25 (1.09, 1.44) |
| Spinal Surg | 0.6% | 0.7% | 0.7% | 0.7% | 0.6% | 0.6% | 0.6% | 0.4% | 0.6% | 0.82 (0.65, 1.05) | 0.99 (0.79, 1.24) | 1.13 (0.91, 1.41) | 1.01 (0.81, 1.26) | 1.08 (0.85, 1.37) | 0.79 (0.62, 1.02) | 1.24 (0.99, 1.54) | 0.91 (0.74, 1.12) | 0.90 (0.71, 1.14) | 1.06 (0.84, 1.33) | 0.64 (0.51, 0.80) | 0.80 (0.63, 1.03) |
| Medicare Advantage | | | | | | | | | | | | | | | | | | | | | |
| At least one | 52167 | 56931 | 31277 | 56383 | 61771 | 33312 | 63274 | 71268 | 36711 | | | | | | | | | | | | |
| AC | 10.6% | 14.2% | 14.4% | 8.8% | 11.8% | 11.6% | 11.1% | 14.3% | 14.1% | 0.73 (0.71, 0.76) | 0.99 (0.96, 1.02) | 0.76 (0.73, 0.79) | 1.02 (0.98, 1.06) | 0.78 (0.76, 0.81) | 1.01 (0.98, 1.04) | 0.83 (0.80, 0.86) | 0.83 (0.80, 0.85) | 0.80 (0.77, 0.84) | 1.05 (1.01, 1.08) | 1.00 (0.98, 1.03) | 0.98 (0.95, 1.02) |
| MT | 5.0% | 6.8% | 8.2% | 4.1% | 5.4% | 6.3% | 4.6% | 6.6% | 7.4% | 0.62 (0.58, 0.65) | 0.82 (0.79, 0.87) | 0.65 (0.62, 0.69) | 0.87 (0.82, 0.91) | 0.63 (0.60, 0.66) | 0.90 (0.86, 0.94) | 0.81 (0.77, 0.86) | 0.80 (0.77, 0.84) | 0.76 (0.72, 0.81) | 0.92 (0.88, 0.97) | 0.98 (0.94, 1.02) | 0.90 (0.85, 0.95) |
| CMT | 9.1% | 12.8% | 9.9% | 8.3% | 11.5% | 8.9% | 9.6% | 11.7% | 9.9% | 0.92 (0.88, 0.96) | 1.29 (1.24, 1.34) | 0.93 (0.89, 0.98) | 1.29 (1.24, 1.35) | 0.97 (0.93, 1.00) | 1.17 (1.13, 1.22) | 0.91 (0.87, 0.94) | 0.90 (0.87, 0.92) | 0.89 (0.85, 0.94) | 1.05 (1.01, 1.09) | 0.91 (0.88, 0.94) | 1.00 (0.95, 1.05) |
| PT | 2.3% | 2.0% | 2.9% | 1.7% | 1.5% | 2.0% | 1.8% | 1.5% | 2.2% | 0.79 (0.72, 0.86) | 0.69 (0.64, 0.76) | 0.88 (0.80, 0.97) | 0.79 (0.71, 0.87) | 0.86 (0.78, 0.94) | 0.72 (0.65, 0.78) | 0.77 (0.70, 0.83) | 0.78 (0.71, 0.85) | 0.69 (0.62, 0.76) | 0.82 (0.76, 0.89) | 0.78 (0.71, 0.84) | 0.75 (0.68, 0.83) |
| Acu | 0.0% | 0.1% | 1.0% | 0.1% | 0.1% | 0.6% | 0.1% | 0.2% | 1.0% | 0.05 (0.03, 0.07) | 0.07 (0.05, 0.10) | 0.17 (0.13, 0.22) | 0.22 (0.17, 0.29) | 0.13 (0.10, 0.16) | 0.16 (0.13, 0.20) | 2.37 (1.47, 3.84) | 1.93 (1.33, 2.80) | 0.64 (0.54, 0.76) | 2.90 (1.83, 4.61) | 2.30 (1.61, 3.28) | 1.04 (0.89, 1.21) |
| OMT | 0.2% | 0.4% | 0.5% | 0.1% | 0.3% | 0.5% | 0.2% | 0.2% | 0.4% | 0.45 (0.35, 0.57) | 0.76 (0.62, 0.94) | 0.30 (0.23, 0.39) | 0.65 (0.53, 0.81) | 0.44 (0.34, 0.55) | 0.57 (0.46, 0.71) | 0.66 (0.49, 0.88) | 0.85 (0.70, 1.03) | 0.99 (0.79, 1.24) | 0.85 (0.65, 1.10) | 0.66 (0.53, 0.80) | 0.87 (0.69, 1.09) |
| Image - Rad | 35.1% | 35.3% | 32.0% | 31.7% | 32.6% | 30.1% | 36.0% | 37.4% | 35.8% | 1.10 (1.07, 1.12) | 1.10 (1.08, 1.12) | 1.05 (1.03, 1.07) | 1.08 (1.06, 1.11) | 1.01 (0.99, 1.02) | 1.04 (1.03, 1.06) | 0.90 (0.89, 0.92) | 0.92 (0.91, 0.94) | 0.94 (0.92, 0.96) | 1.03 (1.01, 1.04) | 1.06 (1.05, 1.08) | 1.12 (1.10, 1.14) |
| Rx - NSAID | 21.9% | 17.9% | 21.5% | 22.4% | 18.2% | 20.8% | 21.3% | 16.8% | 19.0% | 1.02 (0.99, 1.05) | 0.83 (0.81, 0.85) | 1.08 (1.05, 1.11) | 0.88 (0.85, 0.90) | 1.12 (1.09, 1.15) | 0.88 (0.86, 0.90) | 1.02 (1.00, 1.04) | 1.02 (1.00, 1.05) | 0.97 (0.94, 1.00) | 0.97 (0.95, 0.99) | 0.94 (0.92, 0.96) | 0.89 (0.86, 0.91) |
| Image - MRI | 13.9% | 14.5% | 13.4% | 13.5% | 14.1% | 13.4% | 16.3% | 17.6% | 16.4% | 1.04 (1.00, 1.08) | 1.08 (1.05, 1.12) | 1.01 (0.98, 1.05) | 1.05 (1.02, 1.09) | 1.00 (0.97, 1.02) | 1.07 (1.04, 1.10) | 0.98 (0.95, 1.01) | 0.97 (0.95, 1.00) | 1.00 (0.96, 1.04) | 1.18 (1.15, 1.21) | 1.21 (1.18, 1.24) | 1.23 (1.18, 1.27) |
| Rx - MM Rlx | 15.0% | 11.7% | 12.9% | 14.6% | 12.2% | 13.0% | 15.2% | 12.3% | 12.9% | 1.16 (1.12, 1.20) | 0.90 (0.87, 0.94) | 1.12 (1.08, 1.16) | 0.94 (0.90, 0.97) | 1.18 (1.14, 1.22) | 0.95 (0.92, 0.98) | 0.97 (0.95, 1.00) | 1.04 (1.01, 1.08) | 1.01 (0.97, 1.05) | 1.01 (0.99, 1.04) | 1.05 (1.02, 1.08) | 1.00 (0.96, 1.04) |
| Rx - Steroid | 8.9% | 8.8% | 7.9% | 7.8% | 7.5% | 6.4% | 6.8% | 6.7% | 5.8% | 1.12 (1.07, 1.17) | 1.11 (1.06, 1.16) | 1.21 (1.15, 1.27) | 1.17 (1.11, 1.23) | 1.19 (1.13, 1.25) | 1.16 (1.11, 1.22) | 0.87 (0.84, 0.91) | 0.85 (0.82, 0.88) | 0.81 (0.76, 0.85) | 0.77 (0.74, 0.80) | 0.76 (0.73, 0.79) | 0.73 (0.69, 0.77) |
| Rx - Gaba | 2.3% | 2.2% | 2.7% | 2.1% | 1.9% | 2.7% | 1.8% | 1.6% | 1.9% | 0.85 (0.78, 0.93) | 0.82 (0.75, 0.89) | 0.79 (0.72, 0.86) | 0.72 (0.66, 0.78) | 0.92 (0.84, 1.01) | 0.84 (0.76, 0.92) | 0.91 (0.84, 0.98) | 0.86 (0.79, 0.93) | 0.98 (0.89, 1.08) | 0.77 (0.71, 0.83) | 0.72 (0.67, 0.78) | 0.71 (0.65, 0.79) |
| Rx - Oth | 11.7% | 11.6% | 13.1% | 14.1% | 14.1% | 15.6% | 11.9% | 11.6% | 12.2% | 0.89 (0.86, 0.92) | 0.88 (0.85, 0.92) | 0.90 (0.87, 0.93) | 0.90 (0.87, 0.93) | 0.97 (0.94, 1.00) | 0.95 (0.92, 0.98) | 1.21 (1.17, 1.25) | 1.22 (1.18, 1.25) | 1.19 (1.15, 1.24) | 1.02 (0.99, 1.05) | 1.00 (0.97, 1.03) | 0.93 (0.90, 0.97) |
| Rx - Opioid | 19.6% | 13.6% | 16.6% | 19.0% | 14.1% | 16.2% | 17.8% | 13.5% | 14.6% | 1.18 (1.14, 1.22) | 0.82 (0.79, 0.84) | 1.17 (1.14, 1.20) | 0.87 (0.84, 0.90) | 1.22 (1.18, 1.25) | 0.93 (0.90, 0.96) | 0.97 (0.95, 0.99) | 1.04 (1.01, 1.07) | 0.98 (0.94, 1.01) | 0.91 (0.89, 0.93) | 1.00 (0.97, 1.03) | 0.88 (0.85, 0.91) |
| Spinal Inj | 8.4% | 8.7% | 7.4% | 8.1% | 8.4% | 6.8% | 9.0% | 9.2% | 7.9% | 1.14 (1.09, 1.20) | 1.18 (1.13, 1.24) | 1.19 (1.14, 1.25) | 1.24 (1.18, 1.30) | 1.14 (1.09, 1.19) | 1.16 (1.12, 1.21) | 0.96 (0.92, 1.00) | 0.96 (0.93, 1.00) | 0.92 (0.87, 0.97) | 1.07 (1.03, 1.11) | 1.06 (1.02, 1.10) | 1.08 (1.02, 1.13) |
| Image - CT | 6.5% | 6.0% | 5.5% | 6.5% | 6.2% | 5.5% | 7.5% | 7.0% | 6.4% | 1.19 (1.12, 1.26) | 1.10 (1.04, 1.17) | 1.18 (1.12, 1.25) | 1.12 (1.06, 1.18) | 1.16 (1.10, 1.22) | 1.09 (1.04, 1.14) | 1.00 (0.95, 1.04) | 1.02 (0.98, 1.07) | 1.00 (0.94, 1.07) | 1.14 (1.10, 1.19) | 1.16 (1.11, 1.21) | 1.17 (1.11, 1.25) |
| Spinal Surg | 0.6% | 0.7% | 0.6% | 0.6% | 0.6% | 0.7% | 0.6% | 0.6% | 0.5% | 1.09 (0.91, 1.31) | 1.20 (1.01, 1.44) | 0.87 (0.74, 1.04) | 0.96 (0.82, 1.14) | 1.09 (0.91, 1.30) | 1.09 (0.92, 1.30) | 0.94 (0.80, 1.09) | 0.93 (0.81, 1.07) | 1.17 (0.96, 1.42) | 0.90 (0.77, 1.05) | 0.81 (0.71, 0.94) | 0.90 (0.73, 1.10) |

Pre=Pre-COVID period, Early=first 12 months post-COVID, Late=13-24 months post-COVID

B25=Bottom 25 least stringent States on 4/1/2020 and 4/1/2021, T25=Top 25 most stringent States on 4/1/2020 and 4/1/2021, Mixed=In Bottom 25 on 4/1/2020 and Top 25 on 4/1/2021, or in Top 25 on 4/1/2020 and Bottom 25 on 4/1/2021

AC=active care, MT=manual therapy, CMT=chiropractic manipulative therapy, PT=passive therapy, Acu=acupuncture, OMT=osteopathic manipulative therapy, Image - Rad=plain film radiology, Rx - NSAID=non-steroidal anti-inflammatory prescription, Image - MRI=magnetic resonance imaging scan, Rx - MM Rlx= prescription skeletal muscle relaxant, Rx - Steriod=prescription oral steroid, Rx - Gaba=prescription gabapentin, Rx - Oth=other type of prescription medication, Rx - Opioid=prescription opioid, Spinal Inj=Spinal injection, Image - CT=computed tomography, Spinal Surg=spinal surgical procedure

For risk ratios, cells in red indicate the confidence interval includes 1 and likelihood of receiving a service is not different than the Top 25 or pre-COVID reference
